# Treatment of ANCA-Associated Vasculitis With Avacopan: Results of the ADVOCATE Trial After Readjudication of the Primary Endpoint

**DOI:** 10.64898/2026.09.16.26363171

**Authors:** David Jayne, Peter A. Merkel, Xinyu Tang, Zachary S. Wallace, Casey P. Norris, Nikieia Hayden, Sumita Bhatta, Renato D. Lopes, Amy Stallings

**Affiliations:** Department of Medicine, University of Cambridge, Cambridge, UK; Division of Rheumatology, Department of Medicine, Division of Epidemiology, Department of Biostatistics, Epidemiology, and Informatics, University of Pennsylvania, Philadelphia, PA, USA; Amgen Inc., Thousand Oaks, CA, USA; Duke Clinical Research Institute, Duke University Medical Center, Durham, NC, USA; Duke Clinical Research Institute, Department of Pediatrics, Division of Allergy and Immunology, Duke University School of Medicine, Durham, NC, US

## Abstract

**Background:** The ADVOCATE trial evaluated treatment with avacopan for granulomatosis with polyangiitis (GPA) and microscopic polyangiitis (MPA). Concerns regarding the 2019 primary outcome adjudication procedures in nine participants prompted readjudication in 2026 of primary outcomes for all participants.

**Methods:** Patients with GPA/MPA were randomized 1:1 to prednisone taper or avacopan 30 mg twice daily; participants received treatment with cyclophosphamide or rituximab. An independent, blinded committee readjudicated the primary outcomes of remission at week 26 and sustained remission at week 52. Noninferiority and superiority were established if lower bounds of the 95% confidence interval (CI) for difference in rates were greater than −20.0 and 0.0 percentage points, respectively. Secondary outcomes included glucocorticoid exposure and toxicity, kidney parameters, health-related quality of life, and safety.

**Results:** Among 330 participants, remission at week 26 was achieved by 68.1% and 67.1% in the avacopan and prednisone groups, respectively, in the 2026 readjudication (adjusted difference: 2.2%; 95% CI, −7.5, 11.9), compared with 72.3% and 70.1% in the 2019 adjudication (adjusted difference: 3.4%; 95% CI, −6.0, 12.8). Sustained remission at week 52 was achieved by 61.4% and 52.4%, respectively, in the 2026 readjudication (adjusted difference: 9.8%; 95% CI, −0.3, 19.9), compared with 65.7% and 54.9% in the 2019 adjudication (adjusted difference: 12.5%; 95% CI, 2.6, 22.3). Secondary endpoints favored the avacopan group.

**Conclusions:** Treatment of GPA/MPA with avacopan in combination with rituximab or cyclophosphamide, when compared with a prednisone taper, resulted in similarly high rates of remission and sustained remission, but reductions in glucocorticoid exposure and toxicity.

## INTRODUCTION

Anti-neutrophil cytoplasmic antibody (ANCA)–associated vasculitis (AAV) is a heterogeneous group of rare systemic autoimmune conditions characterized by necrotizing inflammation of small to medium-sized blood vessels.^1,2^ The two most common types are granulomatosis with polyangiitis (GPA) and microscopic polyangiitis (MPA).^3,4^ Patients with GPA or MPA often experience life-and organ-threatening manifestations, particularly when disease involves the kidneys or lungs.^2,5–7^ While regimens that include long courses of glucocorticoid treatment in combination with rituximab, cyclophosphamide, and/or other immunosuppressants are effective for many patients, important unmet needs remain: long-term immunosuppression is associated with early and delayed risk of severe infections and reduced vaccine efficacy; glucocorticoids are associated with permanent treatment-related toxicities; relapses occur despite therapy to maintain remission and result in irreversible organ damage; improvement in kidney function is often incomplete; and reduced quality of life remains common.^8–16^

In GPA and MPA, innate and adaptive immune responses drive inflammation through ANCA-mediated neutrophil activation and an inflammatory amplification loop, fueled in part by the alternative complement pathway activation dependent on the complement system, including the C5a–C5aR1 interaction.^17,18^ Avacopan is an orally administered, small-molecule C5aR1 antagonist. Treatment guidelines for GPA/MPA recommend induction of remission with immunosuppressive therapies, such as rituximab or cyclophosphamide, in combination with glucocorticoids, followed by prolonged treatment to maintain remission and prevent relapse.^19–21^ Avacopan has been recommended by these international guidelines as part of a strategy to reduce glucocorticoids.^19–22^

In 2026, concerns were raised regarding the original adjudication process used in the ADVOCATE phase 3 trial, which led to retraction of the original ADVOCATE publication.^23–25^ Specifically, although the 2019 readjudication of the primary endpoints for 9 of 331 participants was conducted by a blinded adjudicator after an initial lock of the database and analysis of the primary outcome, the request to conduct these reassessments was prompted by an unblinded quality control process to ensure that the criteria defining remission had been consistently applied. Following this 2019 readjudication, avacopan demonstrated noninferiority for remission at week 26 and superiority for sustained remission at week 52 when compared with a prednisone taper; in the initial analysis of the 2019 data, before reassessment of the outcomes of 9 participants, avacopan was non-inferior to a prednisone taper at both week 26 for remission and week 52 for sustained remission but not superior at week 52.

In response to these concerns, a blinded, independent readjudication of the primary endpoints from ADVOCATE was conducted in 2026 that followed similar processes and parameters as the original adjudication charter. Importantly, there have been no concerns raised regarding the underlying ADVOCATE data which were used to assess secondary outcomes, including kidney parameters, health-related quality of life, glucocorticoid toxicity, or adverse events. Presented here are the results of a full re-analysis of the primary endpoints based on the 2026 readjudication, in addition to results based on the secondary outcomes.

## METHODS

### Study Design

The trial design has been described previously.^26^ ADVOCATE (NCT02994927) was a phase 3, multicenter, randomized, double-blind, double-dummy, active-controlled trial conducted at 143 centers between March 15, 2017, and November 1, 2019. Patients were randomized 1:1 to receive either avacopan-matching placebo and a prednisone taper (prednisone taper group) or avacopan 30 mg twice daily and prednisone-matching placebo (avacopan group). Both treatment groups received background immunosuppressive induction therapy with cyclophosphamide (followed by azathioprine or mycophenolate mofetil) or rituximab, assigned at the discretion of the investigator (additional details in **Supplementary Material** and **Supplementary Table S1**).

Randomization was stratified according to vasculitis disease status (newly diagnosed or relapsing), ANCA status (anti-proteinase 3 antibody [PR3-ANCA] positive or anti-myeloperoxidase antibody [MPO-ANCA] positive), and non-glucocorticoid immunosuppressive treatment (cyclophosphamide or rituximab). Randomization was performed centrally via an interactive web-response system and minimization algorithm using the stratification factors.^27^

Immunosuppressive and glucocorticoid therapy was protocolized for 52 weeks. For patients in the prednisone taper group, prednisone (or matched placebo) was tapered over the first 20 weeks, with complete discontinuation by week 21 (**Supplementary Table S2**). For patients in the avacopan group who received glucocorticoids during the screening period, the glucocorticoid dose was reduced to ≤20 mg/day prednisone equivalent on day 1 and tapered to 0 mg/day by the end of week 4. Non–study-supplied glucocorticoids were permitted as premedication for rituximab to reduce hypersensitivity reactions, treat persistent or worsening vasculitis, or relapse of vasculitis, as well as for non-vasculitis reasons, such as adrenal insufficiency, though their use was to be minimized as much as possible. No rituximab was given beyond the first 4 weeks. Prophylactic therapy for infection, including for *Pneumocystis jirovecii* pneumonia, was required.

The trial was performed in accordance with the principles of the Declaration of Helsinki and Good Clinical Practice guidelines. Ethics committees and institutional review boards at participating sites approved the research protocol. All patients or a parent or guardian gave written informed consent before entry. ChemoCentryx sponsored the trial and provided trial medication. Medpace conducted the trial with guidance from ChemoCentryx.

### Eligibility Criteria

The ADVOCATE trial enrolled patients aged over 12 years with newly diagnosed or relapsing GPA or MPA, according to the Chapel Hill Consensus Conference definitions, for which treatment with cyclophosphamide or rituximab was indicated, who had new or historic PR3-ANCA or MPO-ANCA positivity, an estimated glomerular filtration rate (eGFR) of at least 15 mL/min/1.73 m^2^, and at least 1 major or 3 non-major items or at least 2 renal items of hematuria and proteinuria on the Birmingham Vasculitis Activity Score (BVAS), version 3. Non-glucocorticoid immunosuppressant drugs used at the time of screening were withdrawn prior to randomization. Patients were excluded if they had received >3 g of intravenous glucocorticoids within 4 weeks or an oral daily dose of glucocorticoids >10 mg prednisone equivalent for >6 weeks continuously before screening.

### Assessments

The primary efficacy endpoints were disease remission at week 26 (defined as a BVAS=0 and no glucocorticoids for treatment of AAV within 4 weeks prior to week 26) and sustained disease remission at week 52 (defined as a BVAS=0 at weeks 26 and 52, no glucocorticoids for the treatment of AAV within 4 weeks prior to weeks 26 and 52, and no relapse between weeks 26 and 52). Patients were not considered to be in remission if they had a BVAS>0 during the 4 weeks prior to week 26 (if collected for an unscheduled assessment). Relapse was defined as worsening of disease after previous achievement of a BVAS=0 involving: 1 or 2 minor BVAS items recorded at 2 consecutive study visits, 1 or more major BVAS items, or 3 or more minor BVAS items.

Secondary endpoints as reported in the 2019 analysis are included here. The secondary efficacy endpoints were change from baseline in eGFR, urinary albumin:creatinine ratio (UACR), glucocorticoid-induced toxic effects according to the Glucocorticoid Toxicity Index (GTI) during the first 26 weeks (measured by both the Cumulative Worsening Score [GTI-CWS], which ranges from 0 to 410, and the Aggregate Improvement Score [GTI-AIS], which ranges from –317 to 410; on both scales, higher scores indicate greater severity of toxic effects; **Supplementary Table S3**),^28,29^ and change from baseline in health-related quality of life assessed with the 36-Item Short Form Health Survey (SF-36), version 2, and the EuroQol Group 5-Dimensions 5-Level Questionnaire (EQ-5D-5L) (range, 0 to 100 for both, with higher scores indicating better quality of life).^30,31^

### Independent, Retrospective Readjudication Analysis

In the ADVOCATE trial, remission, sustained remission, and relapse were assessed by an independent adjudication committee (the 2019 adjudication), whose members were blinded to the treatment assignment, based on the data collected throughout the trial.

To address concerns raised regarding the readjudication of the 9 participants in the 2019 analysis, a blinded, independent readjudication (the 2026 readjudication) of the primary endpoints in the ADVOCATE trial was performed by the Duke Clinical Research Institute (DCRI) Clinical Events Classification (CEC) group.^32^

The 2026 readjudication committee was instructed to adjudicate cases in accordance with an Adjudication Charter (**Supplementary Material**) that followed processes and assessment parameters similar to those of the original ADVOCATE Adjudication Committee Charter. The scope of the readjudication included BVAS, Vasculitis Damage Index (VDI), relapse, and remission at weeks 26 through sustained remission at week 52, and unscheduled and early termination (ET) visits occurring during this period. Amgen commissioned the 2026 readjudication but had no role in the group’s review of data or the determination of remission status.

The DCRI CEC group is an independent academic clinical events adjudication group with extensive experience in endpoint adjudication across all phases of clinical trials and a broad range of therapeutic areas. Physicians serving on the 2026 readjudication committee were selected based on clinical expertise in rheumatology, immunology, nephrology, and cardiology, and knowledge of clinical trial methodology and CEC processes. Adjudicators who participated in the initial ADVOCATE trial were ineligible to serve as 2026 adjudicators. All adjudicators were trained on the protocol, event definitions, existing source documentation, adjudication forms, and operational processes. All adjudicators were trained and certified on BVAS and VDI assessment criteria, through a process proctored and measured by Oxford University. Adjudications were performed using secure systems with established quality control procedures to support the generation of high-quality clinical endpoint data.^32^

The 2026 readjudication committee used case packages with identical data as those provided to the original (2019) adjudication committee, with no modification to the source data. The 2026 readjudication committee members did not have any knowledge of patient-level adjudication decisions made by the original adjudication committee and did not have the opportunity to raise queries with trial sites. An attestation was obtained from the adjudicators confirming that they were not inadvertently unblinded to the subject identifiers or 2019 results of the nine participants based on information that was made public.

### Statistical Analysis

A sample size of 150 participants per group was calculated to provide the trial with at least 90% power to demonstrate noninferiority of avacopan to prednisone taper with respect to the primary endpoint of remission at week 26, assuming a noninferiority margin of −20 percentage points and an incidence of remission in the prednisone taper group of 60%. This sample size also provided 90% power to detect approximately 18% superiority in the proportion of participants achieving remission at week 26.

A supplemental statistical analysis plan was developed that replicated the analytic plan for the primary and select secondary endpoints as detailed in the final 2019 statistical analysis plan. The primary efficacy analyses were conducted in the intention-to-treat population, defined as all randomized participants who received at least one dose of trial medication. For each primary endpoint, the difference in the proportion of participants achieving remission was estimated using inverse-variance stratum weights (**Supplementary Appendix**), and the confidence interval (CI) for the difference was estimated using the Miettinen–Nurminen method. Participants with missing data at weeks 26 or 52 were considered not to have achieved remission at the corresponding time point. For each primary endpoint, avacopan was considered noninferior to prednisone taper if the lower bound of the two-sided 95% CI for the difference (avacopan minus prednisone taper) was greater than −20 percentage points and the proportion of participants achieving remission in the prednisone taper group was at least 40 percentage points; avacopan was considered superior to prednisone taper if the lower bound of the 95% CI was greater than 0.0 percentage points. To control the overall type I error rate, the two primary endpoints were tested sequentially in the following hierarchical order: noninferiority at week 26, noninferiority at week 52, superiority at week 52, and superiority at week 26. The P-values for noninferiority and superiority were one-sided. Concordance was assessed by evaluating the percent agreement and Cohen’s kappa coefficient between the 2019 and 2026 adjudications. Cohen’s kappa quantifies the level of agreement beyond the expected chance.

Secondary endpoint analyses of continuous variables were performed with the use of mixed-effects models for repeated measures. Least-squares (LS) means, standard errors, and CIs were from models incorporating treatment group, visit, treatment-by-visit interaction, and stratification factors as covariates. Longitudinal measurements from the same patients were considered as repeated-measure units in the model. The Kaplan– Meier method was used to estimate the time to relapse of vasculitis, and the proportionality assumption was upheld. There was no prespecified plan for adjustment of CIs for multiplicity of the secondary endpoints; point estimates and 95% CIs only are presented, and no definite conclusions can be drawn from these data. Prespecified subgroup analyses were performed, but the trial was not powered to make conclusions from these results.

## RESULTS

### Patients

A total of 331 patients were randomized (avacopan group, n=166; prednisone taper group, n=165), but one patient was excluded from the analysis because the patient did not receive prednisone. Baseline demographics and clinical characteristics were reported previously.^24,25^ Baseline characteristics were similar between treatment groups, with similar glucocorticoid doses during the screening period (**Table 1**). In the overall trial population, mean age was 61 years, 43% of patients were PR3-ANCA positive, 57% were MPO-ANCA positive, 69% of patients had newly diagnosed vasculitis, and 81% had kidney involvement. Approximately two-thirds of patients received rituximab, and one-third received cyclophosphamide.

**Table 1.**
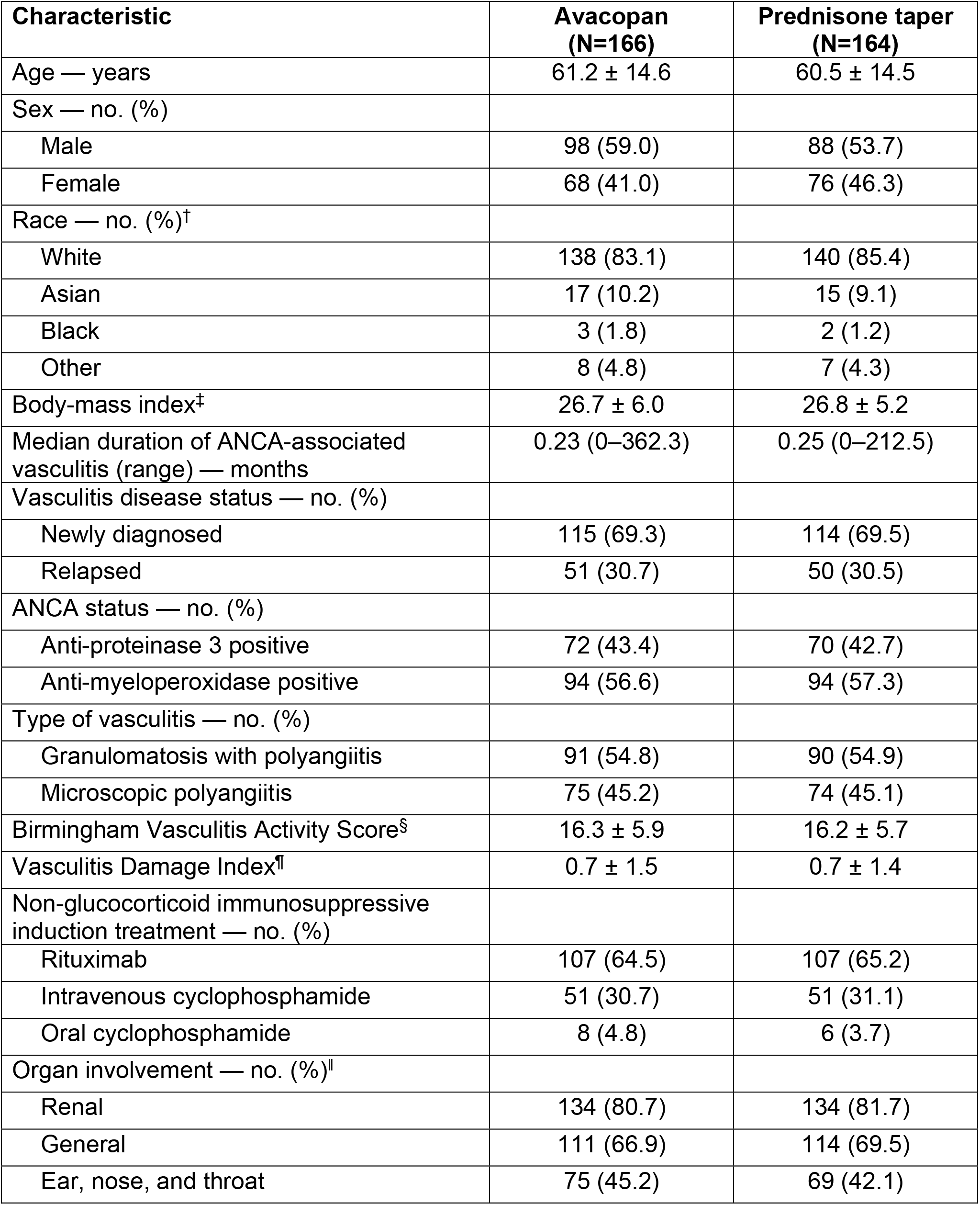
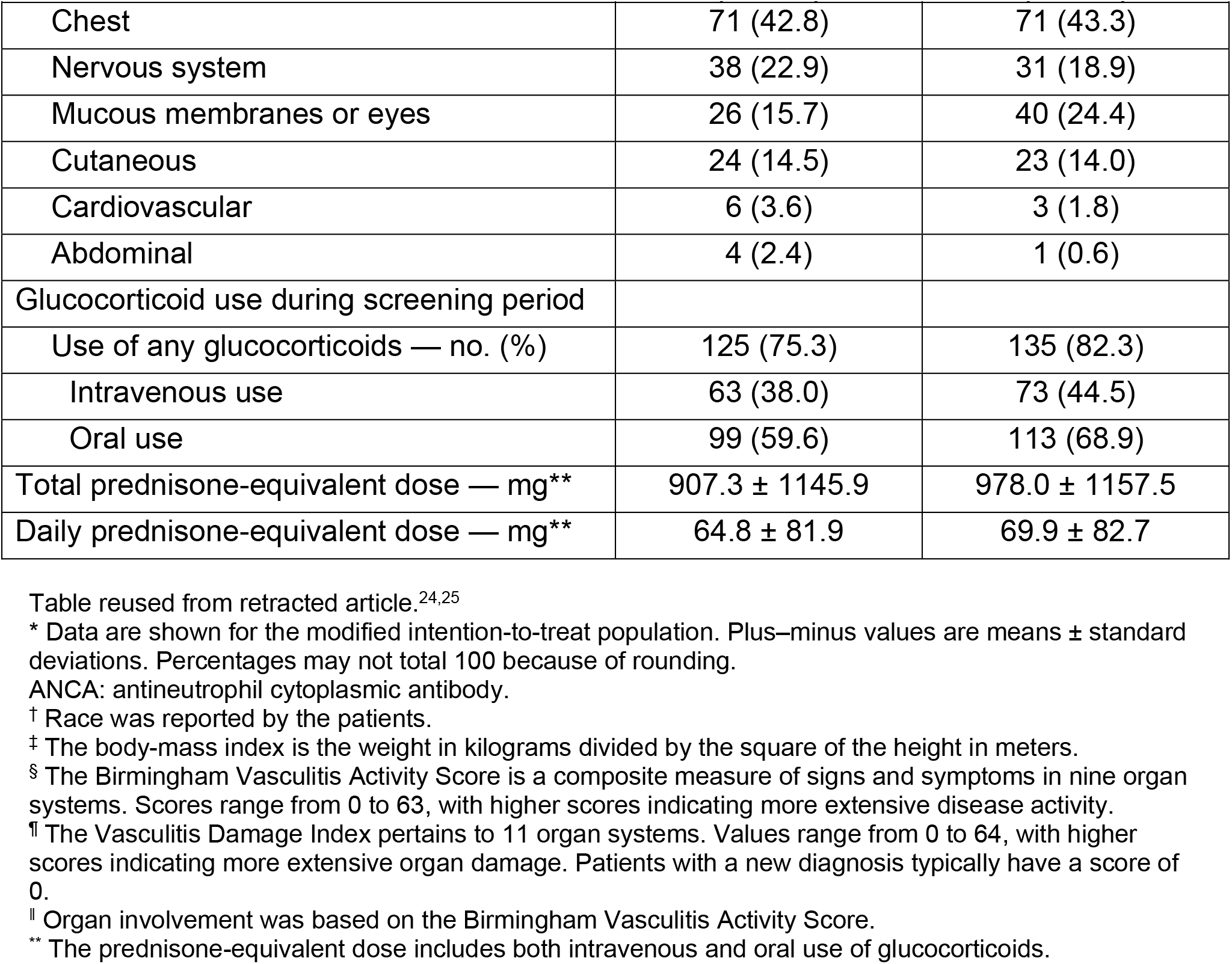
Baseline demographics and clinical characteristics*.

| **Characteristic** | **Avacopan**<br>**(N=166)** | **Prednisone taper**<br>**(N=164)** |
| --- | --- | --- |
| Age — years | 61.2 ± 14.6 | 60.5 ± 14.5 |
| Sex — no. (%) |  |  |
| Male | 98 (59.0) | 88 (53.7) |
| Female | 68 (41.0) | 76 (46.3) |
| Race — no. (%)<sup>†</sup> |  |  |
| White | 138 (83.1) | 140 (85.4) |
| Asian | 17 (10.2) | 15 (9.1) |
| Black | 3 (1.8) | 2 (1.2) |
| Other | 8 (4.8) | 7 (4.3) |
| Body-mass index<sup>‡</sup> | 26.7 ± 6.0 | 26.8 ± 5.2 |
| Median duration of ANCA-associated vasculitis (range) — months | 0.23 (0–362.3) | 0.25 (0–212.5) |
| Vasculitis disease status — no. (%) |  |  |
| Newly diagnosed | 115 (69.3) | 114 (69.5) |
| Relapsed | 51 (30.7) | 50 (30.5) |
| ANCA status — no. (%) |  |  |
| Anti-proteinase 3 positive | 72 (43.4) | 70 (42.7) |
| Anti-myeloperoxidase positive | 94 (56.6) | 94 (57.3) |
| Type of vasculitis — no. (%) |  |  |
| Granulomatosis with polyangiitis | 91 (54.8) | 90 (54.9) |
| Microscopic polyangiitis | 75 (45.2) | 74 (45.1) |
| Birmingham Vasculitis Activity Score<sup>§</sup> | 16.3 ± 5.9 | 16.2 ± 5.7 |
| Vasculitis Damage Index | 0.7 ± 1.5 | 0.7 ± 1.4 |
| Non-glucocorticoid immunosuppressive induction treatment — no. (%) |  |  |
| Rituximab | 107 (64.5) | 107 (65.2) |
| Intravenous cyclophosphamide | 51 (30.7) | 51 (31.1) |
| Oral cyclophosphamide | 8 (4.8) | 6 (3.7) |
| Organ involvement — no. (%) |  |  |
| Renal | 134 (80.7) | 134 (81.7) |
| General | 111 (66.9) | 114 (69.5) |
| Ear, nose, and throat | 75 (45.2) | 69 (42.1) |

| Characteristic | Avacopan<br>(N=166) | Prednisone taper<br>(N=164) |
| --- | --- | --- |
| Chest | 71 (42.8) | 71 (43.3) |
| Nervous system | 38 (22.9) | 31 (18.9) |
| Mucous membranes or eyes | 26 (15.7) | 40 (24.4) |
| Cutaneous | 24 (14.5) | 23 (14.0) |
| Cardiovascular | 6 (3.6) | 3 (1.8) |
| Abdominal | 4 (2.4) | 1 (0.6) |
| Glucocorticoid use during screening period |  |  |
| Use of any glucocorticoids — no. (%) | 125 (75.3) | 135 (82.3) |
| Intravenous use | 63 (38.0) | 73 (44.5) |
| Oral use | 99 (59.6) | 113 (68.9) |
| Total prednisone-equivalent dose — mg** | 907.3 ± 1145.9 | 978.0 ± 1157.5 |
| Daily prednisone-equivalent dose — mg** | 64.8 ± 81.9 | 69.9 ± 82.7 |

Table reused from retracted article.<sup>24,25</sup>
\* Data are shown for the modified intention-to-treat population. Plus-minus values are means ± standard deviations. Percentages may not total 100 because of rounding.
ANCA: antineutrophil cytoplasmic antibody.
† Race was reported by the patients.
‡ The body-mass index is the weight in kilograms divided by the square of the height in meters.
§ The Birmingham Vasculitis Activity Score is a composite measure of signs and symptoms in nine organ systems. Scores range from 0 to 63, with higher scores indicating more extensive disease activity.
¶ The Vasculitis Damage Index pertains to 11 organ systems. Values range from 0 to 64, with higher scores indicating more extensive organ damage. Patients with a new diagnosis typically have a score of 0.
|| Organ involvement was based on the Birmingham Vasculitis Activity Score.
\*\* The prednisone-equivalent dose includes both intravenous and oral use of glucocorticoids.

### Primary Endpoints: Remission and Sustained Remission

As described previously, based on the 2019 adjudication, remission was achieved at week 26 by 72.3% (120/166) of patients in the avacopan group and 70.1% (115/164) in the prednisone taper group (estimated common difference [95% CI], 3.4% [−6.0, 12.8]; *P*<0.0001 for noninferiority; *P*=0.2387 for superiority).^24,25^ Sustained remission was achieved at week 52 by 65.7% (109/166) of patients in the avacopan group and 54.9% (90/164) in the prednisone taper group (estimated common difference [95% CI], 12.5% [2.6, 22.3]; *P*<0.0001 for noninferiority; *P*=0.0066 for superiority; **Figure 1**).

**Figure 1.**
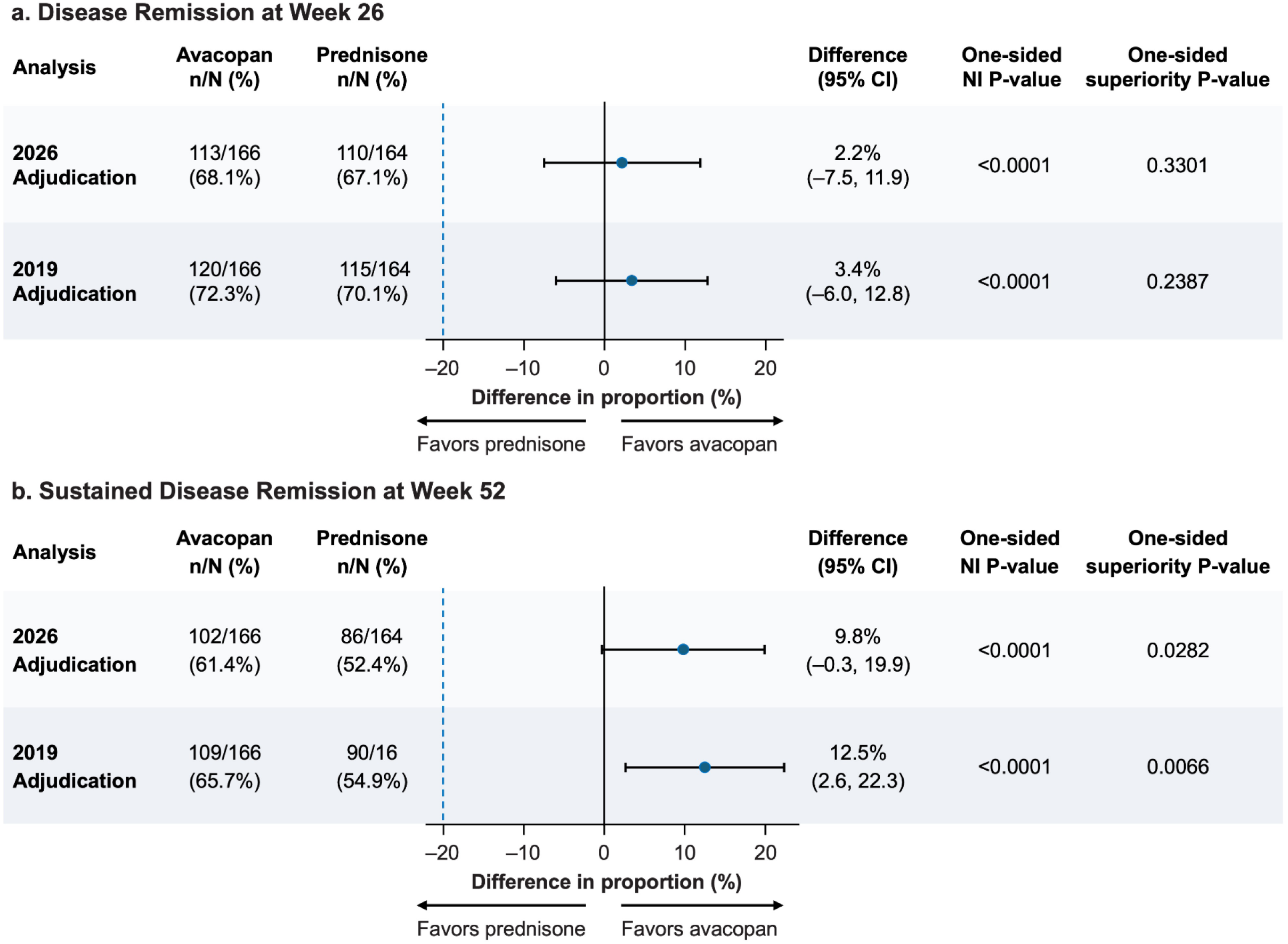
Primary Endpoints: Remission at Week 26 (A) and Sustained Remission at Week 52(B) The 2026 adjudication refers to the retrospective, blinded, independent readjudication of the primary endpoints conducted by the Duke Clinical Research Institute Clinical Events Classification group. The 2019 adjudication refers to the results published in 2021 in the *New England Journal of Medicine* (now retracted).^24,25^ Confidence intervals for treatment proportions are calculated using the Clopper and Pearson Method. Two-sided 95% confidence intervals are calculated for the difference in proportions (avacopan minus prednisone) adjusted for randomization strata (newly diagnosed or relapsed AAV, anti-PR3 or anti-MPO ANCA, and IV rituximab or cyclophosphamide (IV or oral) standard of care treatment) using the stratified summary score test and estimate for the common difference in proportions. Non-inferiority and superiority *P* values are one-sided. AAV, ANCA-associated vasculitis; ANCA, anti-neutrophil cytoplasmic antibody; IV, intravenous; MPO, myeloperoxidase; NI, non-inferiority; PR3, proteinase-3.

Based on the 2026 readjudication, remission was achieved at week 26 by 68.1% (113/166) of patients in the avacopan group and 67.1% (110/164) in the prednisone taper group (estimated common difference [95% CI], 2.2% [−7.5, 11.9]; *P*<0.0001 for noninferiority; *P*=0.3301 for superiority) (**Figure 1**). Sustained remission was achieved at week 52 by 61.4% (102/166) of patients in the avacopan group and 52.4% (86/164) in the prednisone taper group (estimated common difference [95% CI], 9.8% [−0.3, 19.9]; *P*<0.0001 for noninferiority; *P*=0.0282 for superiority; **Figure 1**).

Remission results for per-protocol analyses based on the 2026 readjudication are shown in **Supplementary Table S4.** Achievement of remission and sustained remission was generally consistent across subgroups of background immunosuppressive therapy, ANCA type, vasculitis disease status, and kidney involvement in the 2026 adjudication (**Supplementary Figure S1**), which was consistent with the findings from the 2019 adjudication (**Supplementary Tables S5 and S6**).

### Concordance Between the 2019 and 2026 Adjudications

Concordance between the 2019 adjudication and the 2026 adjudication analyses was 95.2% (Cohen’s kappa 0.89) for the assessment of remission at week 26 and 93.6% (Cohen’s kappa 0.87) for sustained remission at week 52. The factors contributing to differences between the 2019 and 2026 adjudication analyses included differences in the adjudication committees’ assessments of BVAS, reasons for glucocorticoid use in the 4 weeks preceding weeks 26 and/or 52, and occurrences of relapse between weeks 26 and 52 (**Figure 2**). Because the 2026 readjudication reassessed the primary outcomes for all participants, outcome classifications could have changed for any participant between the 2019 and 2026 analyses. **Supplementary Table S7** provides additional detail on the assessments from both adjudications for the nine participants whose 2019 primary outcome adjudication had prompted concerns.

**Figure 2:**
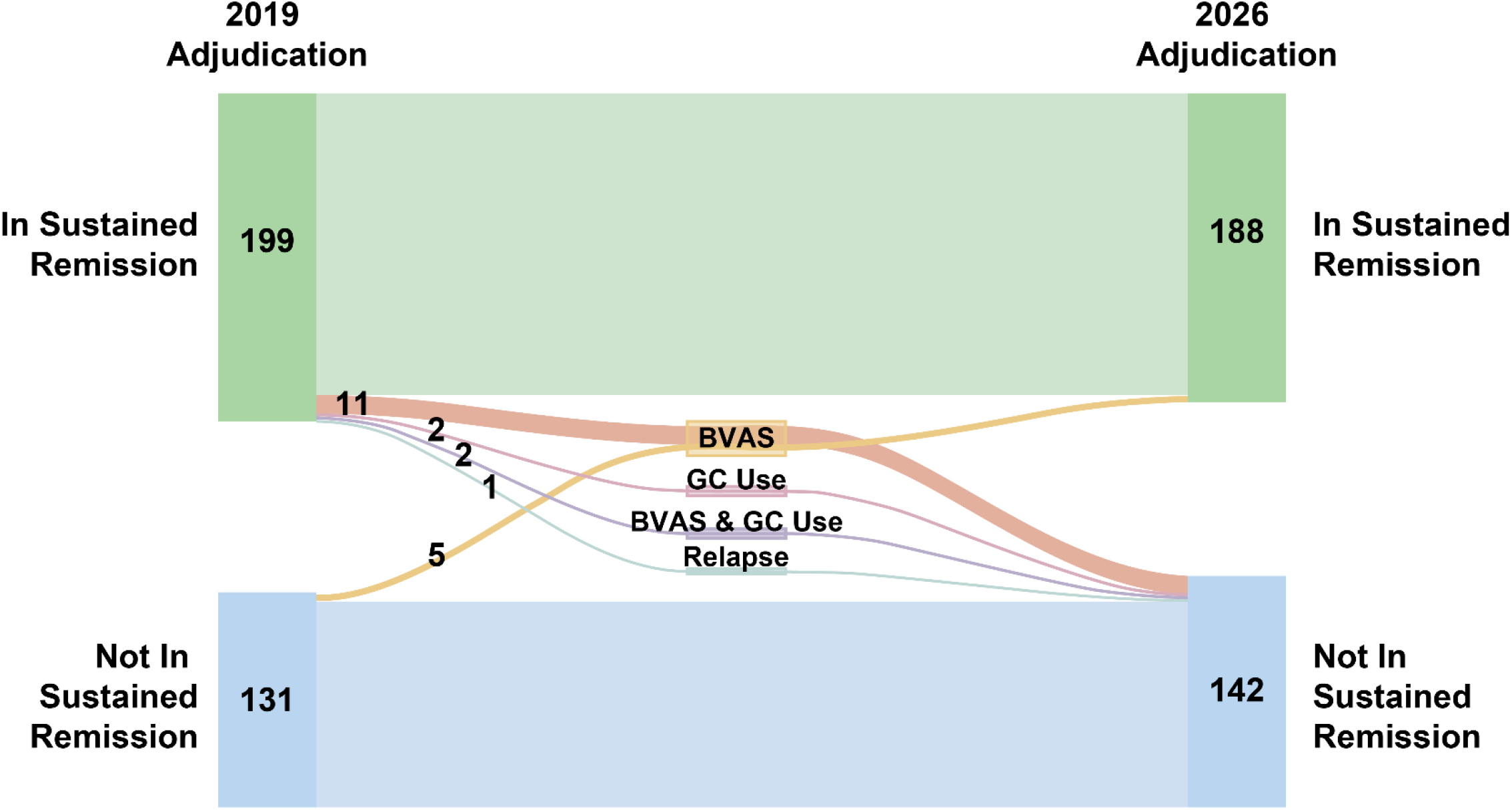
Comparison of Week 52 Sustained Remission Outcomes Across the 2019 and 2026 Adjudications. The factors contributing to differences in the 2019 and 2026 adjudication analyses included differences in the adjudication committees’ assessments of the BVAS score (BVAS), reason for glucocorticoid use in the 4 weeks preceding weeks 26 and/or 52 (GC Use), and occurrence of relapse between weeks 26 and 52 (Relapse). BVAS, Birmingham Vasculitis Activity Score; GC glucocorticoid.

### Disease Relapse

Relapse after previously achieving remission at week 26 occurred in 6.2% (7/113) of patients in the avacopan group and 10.9% (12/110) in the prednisone taper group in the 2026 readjudication (estimated common difference [95% CI], −6.0% [−14.6, 2.6]; *P*=0.08; **Supplementary Table S8**). These findings were similar to those of the 2019 adjudication (**Supplementary Figure S2**).

### Renal Function

At week 52, eGFR was increased relative to the baseline visit in both treatment groups (avacopan: LS mean 7.3 mL/min/1.73 m^2^; prednisone taper: LS mean 4.1 mL/min/1.73 m^2^; difference [95% CI], 3.2 [0.3, 6.1]) (**Figure 3A**). Among patients with baseline eGFR <30 mL/min/1.73 m^2^, eGFR increased by week 52 by an LS mean of 13.7 mL/min/1.73 m^2^ in the avacopan and 8.2 mL/min/1.73 m^2^ in the prednisone taper groups (difference [95% CI], 5.6 mL/min/1.73 m^2^ [1.7, 9.5]; **Supplementary Figure S3**). During the trial, greater decreases were observed in UACR in the avacopan than in the prednisone taper groups at week 4, after which UACR decreased to a similar extent in both groups through week 52 (**Figure 3B**).^24,25^

**Figure 3.**
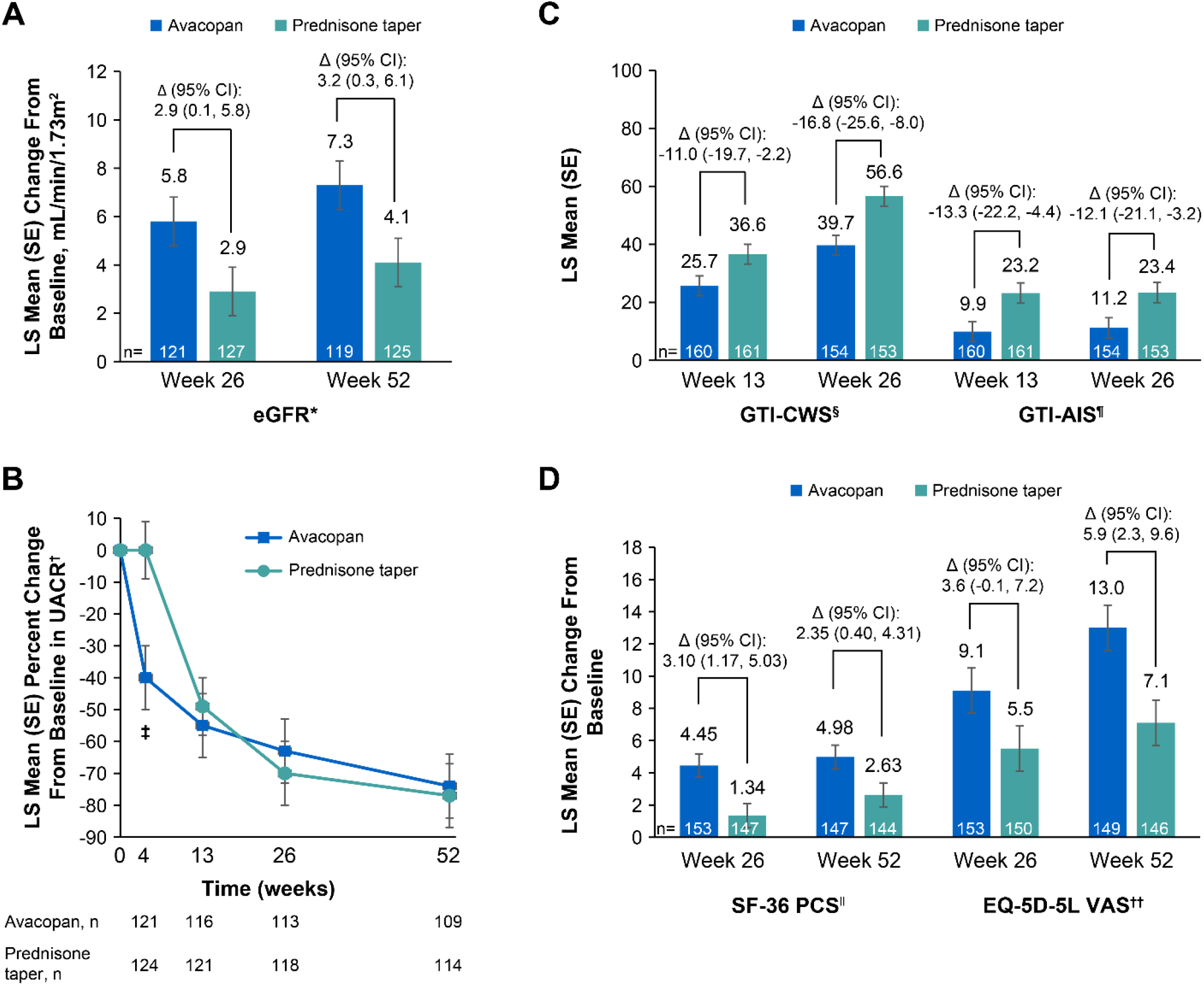
Key secondary endpoints. eGFR (A), UACR (B), GTI-CWS, GTI-AIS (C), SF-36 PCS, and EQ-5D-5L VAS (D). Data previously reported in a retracted article. ^24,25^ *Shown is the estimated glomerular filtration rate (eGFR) in patients with renal disease at baseline on the basis of the BVAS. ^†^Values are for patients with renal disease (on the basis of the BVAS) and a urinary albumin:creatinine ratio (with albumin measured in milligrams and creatinine in grams) of at least 10 at baseline. Percent changes from baseline are based on ratios of geometric means of visit over baseline. ^‡^Indicates where 95% confidence intervals of the differences between the avacopan and prednisone taper arm do not include 0. ^§^The Glucocorticoid Toxicity Index Cumulative Worsening Score (GTI-CWS) ranges from 0 to 410, with higher scores indicating greater severity of toxic effects. ^¶^The Glucocorticoid Toxicity Index Aggregate Improvement Score (GTI-AIS) ranges from –317 to 410, with higher scores indicating greater severity of toxic effects. ^‖^The physical component score on the 36-Item Short Form Health Survey (SF-36), version 2, ranges from 0 to 100, with higher scores indicating better quality of life. ^††^Scores on the visual-analogue scale of the EuroQoL Group 5-Dimensions 5-Level Questionnaire (EQ-5D-5L) range from 0 to 100, with higher scores indicating better quality of life. eGFR, estimated glomerular filtration rate; EQ-5D-5L VAS, EuroQoL Group 5-Dimensions 5-Level Questionnaire visual analog scale; GTI-AIS, Glucocorticoid Toxicity Index Aggregate Improvement Score; GTI-CWS, Glucocorticoid Toxicity Index Cumulative Worsening Score; LS, least-squares; SE, standard error; SF-36 PCS, 36-Item Short Form Health Survey physical component summary; UACR, albumin:creatinine ratio.

### Glucocorticoid use

Participants randomized to avacopan had an 81% reduction in median glucocorticoid exposure. The mean total prednisone-equivalent dose of oral and intravenous glucocorticoids was 1676 mg (equating to 5 mg per patient per day) in the avacopan group and 3847 mg (equating to 13 mg per patient per day) in the prednisone group (**Supplementary Table S9**). Glucocorticoid use according to week is shown in **Supplementary Figures S4 and S5**, and use according to trial period is shown in **Supplementary Table S10**. Non-glucocorticoid immunosuppressants, other than those specified in the protocol, for worsening of vasculitis or relapses, were used in 29 patients (17.5%) in the avacopan group and in 36 patients (22.0%) in the prednisone group (**Supplementary Table S10**).

### Glucocorticoid Toxicity Index

The LS mean for the GTI-CWS at week 26 was lower in the avacopan group than in the prednisone taper group (39.7 vs 56.6; difference [95% CI], −16.8 [−25.6, −8.0]) (**Figure 3C**).^24,25^ Similarly, LS mean GTI-AIS was lower at week 26 in the avacopan group than in the prednisone taper group (11.2 vs 23.4; difference [95% CI], −12.1 [−21.1, −3.2]).

### Health-Related Quality of Life

Numerically greater improvements in health-related quality of life, as measured by SF-36 physical component summary and EQ-5D-5L visual analog scale, were observed in the avacopan versus the prednisone taper groups at weeks 26 and 52 (**Figure 3D, Supplementary Figures S6 and S7; Supplementary Table S8**).

### Safety

There were 116 serious adverse events (AEs) in the avacopan group and 166 in the prednisone taper treatment group, with similar percentages of patients experiencing a serious AE in each group (avacopan, 42.2%; prednisone taper, 45.1%) (**Table 2**).^24,25^ The most common serious AE was worsening vasculitis (avacopan, 10.2%; prednisone taper, 14.0%). Rates of discontinuation due to AEs were similar between groups (avacopan, 15.7%; prednisone taper, 17.7%). There were 2 deaths in the avacopan (worsening vasculitis and pneumonia) and 4 in the prednisone taper groups (generalized fungal infection, infectious pleural effusion, acute myocardial infarction, and death of unknown cause). There were 1 fatal infection and 1 life-threatening infection in the avacopan group and 2 of each in the prednisone taper group. Serious infections/serious opportunistic infections occurred in 13.3%/3.6% of participants in the avacopan group and 15.2%/6.7% in the prednisone taper group (**Supplementary Table S11**). Serious AEs of abnormalities in liver-function testing occurred in 9 (5.4%) patients in the avacopan group and 6 (3.7%) in the prednisone taper treatment group. All events resolved following withdrawal of trial medication and other potentially hepatotoxic drugs, including trimethoprim–sulfamethoxazole. AEs potentially related to glucocorticoids (**Supplementary Table S12**) occurred in 66.3% of participants in the avacopan group and 80.5% in the prednisone taper group.^24,25^

**Table 2.** Safety Results*.

| **Event** | **Avacopan**<br>**(N=166)** | **Prednisone taper**<br>**(N=164)** |
| --- | --- | --- |
| Any adverse event |  |  |
| No. of patients (%) | 164 (98.8) | 161 (98.2) |
| No. of events | 1779 | 2139 |
| Severe adverse event<sup>†</sup> |  |  |
| No. of patients (%) | 39 (23.5) | 41 (25.0) |
| No. of events | 71 | 94 |
| Life-threatening adverse event |  |  |
| No. of patients (%) | 8 (4.8) | 14 (8.5) |
| No. of events | 8 | 22 |
| Death – no. (%) | 2 (1.2) | 4 (2.4) |
| Any serious adverse event<sup>‡</sup> |  |  |
| No. of patients (%) | 70 (42.2) | 74 (45.1) |
| No. of events | 116 | 166 |
| Any serious adverse event related to vasculitis worsening<sup>§</sup> |  |  |
| No. of patients (%) | 17 (10.2) | 23 (14.0) |
| No. of events | 18 | 36 |
| Any serious adverse event not related to vasculitis worsening |  |  |
| No. of patients (%) | 62 (37.3) | 64 (39.0) |
| No. of events | 98 | 130 |
| Discontinuation of trial medication due to adverse event – no. (%) | 26 (15.7) | 29 (17.7) |
| Any infection |  |  |
| No. of patients (%) | 113 (68.1) | 124 (75.6) |
| No. of events | 233 | 291 |
| Any serious infection<sup>¶</sup> |  |  |
| No. of patients (%) | 22 (13.3) | 25 (15.2) |
| No. of events | 25 | 31 |
| Any serious opportunistic infection – no. (%) | 6 (3.6) | 11 (6.7) |
| Death due to infection – no. (%) | 1 (0.6) | 2 (1.2) |
| Life-threatening infection – no. (%) | 1 (0.6) | 2 (1.2) |

| Event | Avacopan<br>(N=166) | Prednisone taper<br>(N=164) |
| --- | --- | --- |
| Serious adverse event of abnormality on liver-function testing – no. (%) | 9 (5.4) | 6 (3.7) |
| Any adverse event potentially related to glucocorticoids – no. (%)** | 110 (66.3) | 132 (80.5) |
| Cardiovascular | 72 (43.4) | 85 (51.8) |
| Infectious | 22 (13.3) | 25 (15.2) |
| Gastrointestinal | 3 (1.8) | 4 (2.4) |
| Psychological | 27 (16.3) | 39 (23.8) |
| Endocrine or metabolic | 23 (13.9) | 48 (29.3) |
| Dermatologic | 14 (8.4) | 28 (17.1) |
| Musculoskeletal | 19 (11.4) | 21 (12.8) |
| Ophthalmologic | 7 (4.2) | 12 (7.3) |
| Any adverse event potentially related to glucocorticoids as assessed by the investigators – no. (%) | 107 (64.5) | 131 (79.9) |
| Any serious adverse event potentially related to prednisone as assessed by the investigators – no. (%) | 11 (6.6) | 24 (14.6) |

Table reused from retracted article.<sup>24,25</sup>
\* Incidence is expressed as number and percentage of patients having at least one event.
† Severe adverse events were defined as those events that caused an inability of a patient to carry out usual activities.
‡ Serious adverse events were defined as any adverse event that resulted in death, was immediately life-threatening, required or prolonged hospitalization, resulted in persistent or clinically significant disability or incapacity, was a birth defect, or was an important event that might jeopardize the patient or might have required intervention to prevent any of the above.
§ Data are for patients who had a serious adverse event of ANCA-positive vasculitis (worsening), granulomatosis with polyangiitis (worsening), or microscopic polyangiitis (worsening).
¶ All serious infections are summarized in Table S11.
|| The life-threatening infections were not the same as the fatal infections. One patient in the prednisone group had sepsis and another had bacteremia and meningitis. One patient in the avacopan group, who also received two rituximab infusions before the event, had hepatitis C reactivation during the trial drug-free follow-up period.
\*\* Predefined *Medical Definition for Regulatory Activities* preferred terms based on the European League against Rheumatism search criteria<sup>53</sup> were included for each cluster (see Table S12 for included terms).

## DISCUSSION

Patients living with GPA and MPA have a serious condition that can be life-threatening and have few treatment options. Across recent clinical trials, a substantial portion of participants failed to achieve remission, highlighting the critical unmet need for new treatments.^33,34^ This reanalysis of the results of the ADVOCATE trial based on the 2026 readjudication of the primary endpoints demonstrated that avacopan, when used in combination with rituximab or cyclophosphamide for induction of remission, was associated with high rates of remission GPA and MPA at week 26 and sustained remission at week 52, utilizing a treatment strategy that substantially reduced glucocorticoid exposure and associated toxicity in participants with GPA and MPA. There was a high degree of concordance regarding the primary endpoints assessment between the 2019 and 2026 adjudication committees.

As in the 2019 analysis, the lower bound of the CIs in the 2026 re-analysis was well above the pre-specified noninferiority margin of −20%, providing high confidence in the noninferiority results at weeks 26 and 52. Although statistical significance, which required a p-value of <0.025, for superiority of sustained remission at week 52 was not achieved with avacopan in the 2026 readjudication, the between-group difference was 9.8% (95% CI, −0.3, 19.9; P=0.0282), a strong numerical trend, which was also consistent with that observed in the 2019 adjudication (12.5%; 95% CI, 2.6, 22.3; P=0.007).

Blinded endpoint adjudication committees are commonly used in clinical trials to improve the consistency and objectivity of endpoint assessment, particularly when endpoint determination requires clinical judgment.^35^ As done in ADVOCATE, adjudication committees apply prespecified criteria in accordance with an adjudication charter, and predefined procedures exist to resolve differences in interpretation among reviewers. Consequently, some differences in assessments between independent adjudication committees might occur. BVAS is a semi-objective endpoint measure that is subject to differing interpretation between scorers, which may introduce additional variability in assessments.

Unlike the 2019 adjudication committee, the 2026 adjudication committee was unable to query sites if members had questions regarding investigator assessments. To mitigate this limitation, the 2026 adjudication committee was provided with the query log from the 2019 adjudication committee. Despite the inherent variability of endpoint adjudication and retrospective nature of the 2026 readjudication, concordance between the original and 2026 adjudications was high, supporting the reliability of the findings.

Much of the discrepancy between the 2019 and 2026 adjudications was related to the BVAS assessments, with 11 participants reclassified from remission status to no remission status (i.e. from BVAS=0 to BVAS ≥1). Notably, of the disease status of the nine patients who underwent readjudication after the initial 2019 analysis for the week 52 end-point, only one differed between the two adjudications. Because the assessments were post-baseline, the BVAS items would either have been new, reflecting relapse of vasculitis, or persisting from the preceding evaluation. Adjudication of such persisting items on the BVAS is subject to variation because investigators and adjudicators may differ on whether such recurring items represent persistent active vasculitis or are due to damage from the disease. It is important to stress that for both 2019 and 2026 adjudications, the adjudicators were blinded to treatment allocation, so any systematic biases should be balanced between treatment groups.

There remains a high unmet need for therapeutic options for treating GPA and MPA. Despite advances over the last several decades, patients with GPA and MPA continue to experience suboptimal outcomes, including relapses, chronic kidney disease, treatment-related toxicities, including those from the use of glucocorticoids and long-term immunosuppression, reduced quality of life, and markedly increased mortality risk.^36^ Glucocorticoids have been a cornerstone of treatment of GPA and MPA, but cumulative glucocorticoid exposure is associated with substantial toxicities that contribute to much of the morbidity that patients with GPA and MPA experience.^21,37^

The findings in the ADVOCATE trial support the important role that avacopan can serve for patients with GPA and MPA by helping them achieve high rates of remission and sustained remission utilizing a strategy that minimizes glucocorticoid exposure and glucocorticoid toxicity. Furthermore, improvements in kidney outcomes and quality of life observed in ADVOCATE, along with a growing body of real-world evidence, including comparative effectiveness studies, contribute to the understanding of the effectiveness of avacopan.^38–43^

In this trial, there was a lower rate of infections and there were fewer AEs associated with glucocorticoid use in the avacopan group than the prednisone taper group, consistent with the reduction in glucocorticoid exposure in the avacopan group.

However, AEs of abnormality on liver-function testing were slightly higher in the avacopan group than in the prednisone taper group (5.4% vs 3.7%). Hepatotoxicity is a known risk associated with treatment with avacopan and is included in product labeling with recommendations for monitoring to mitigate risk.^44^ A detailed analysis of the safety of avacopan across the two Phase II and one Phase III randomized trials was published.^45^ Two Phase II trials and real-world studies support the safety profile of avacopan.^41,46–50^

This study has certain limitations. First, the 2026 readjudication was conducted retrospectively, and while committee members were provided with the query log from the trial, they were unable to query sites with questions related to BVAS scores, glucocorticoid use, or other matters. Second, ADVOCATE was powered to detect an approximate 18% difference in the rates of sustained remission between the two groups and, therefore, was underpowered to demonstrate statistical significance for the smaller treatment difference of 9.8% observed in the 2026 re-analysis. Third, secondary outcomes were not adjusted for multiplicity and should be interpreted cautiously. However, these findings, along with the totality of evidence, including multiple real-world studies, supports the efficacy profile of avacopan.^51,52^

Strengths of this study include the rigorous design of the original ADVOCATE trial, a large, multicenter, randomized, double-blind, double-dummy, active-controlled, phase 3 trial, combined with a fully blinded and independent 2026 readjudication of the primary efficacy endpoints. The 2026 readjudication used prespecified endpoint definitions and procedures aligned with the original adjudication framework with committee members who had not participated in the initial trial, thereby reducing the potential for treatment-related or prior-adjudication bias.

## CONCLUSIONS

The analyses of the ADVOCATE trial, based on the 2026 readjudication of the primary endpoints, reaffirm the noninferiority of avacopan, as compared with a prednisone taper, for remission at week 26 and a consistent treatment effect favoring avacopan for sustained remission at week 52. These findings, combined with real-world evidence and key secondary outcomes from the ADVOCATE trial supporting improvements in kidney outcomes, health-related quality of life, and glucocorticoid-related toxicity, each of which were not impacted by the readjudication process, support that treatment with avacopan is beneficial to patients with GPA/MPA.

## Supporting information

Adjudication Charter

## Data Availability

The data underlying the analyses described in this report are not publicly available. Following completion of the applicable review and governance processes, qualified researchers may submit requests for access to study data for scientifically sound research purposes. Requests will be evaluated by an internal data review committee according to predefined scientific, ethical, legal, and contractual criteria. Access, if approved, will be provided under an appropriate data-sharing agreement and in accordance with applicable laws, regulations, and participant privacy protections.

## ACKNOWLEDGMENTS

Writing and editorial support was funded by Amgen Inc. Rebecca Lane, PhD, of Peloton Advantage, LLC, an OPEN Health company and Venkatesh Taadla, CMPP, of Amgen India, provided medical writing support for this manuscript.

## AUTHOR CONTRIBUTIONS

DJ and PAM conceived the original design of the ADVOCATE trial. SB conceived of the design of the readjudication of the ADVOCATE primary outcomes. AS, XT, CPN, RDL, NH, and SB contributed to the methodology. AS, CPN, RDL, and NH contributed to investigation. XT contributed to software, formal analysis, and data visualization. AS, CPN, RDL, and NH provided supervision, and NH project administration support. AS, CPN, RDL, and NH contributed to data curation. XT and ZSW drafted the original manuscript. DJ, PAM, AS, ZSW, CPN, RDL, NH, and SB reviewed and edited the draft and provided critical feedback. All authors approved the final draft of the manuscript.

