## Supplementary Material for "Treatment of ANCA-Associated Vasculitis With Avacopan: Results of the ADVOCATE Trial After Readjudication of the Primary Endpoint"

#### 1. PARTICIPATING INVESTIGATORS

(Presented by country, with National Coordinating Center followed by participating centers, in alphabetical order by principal investigator.)

**Australia**— National Coordinating Center: Royal Adelaide Hospital, Adelaide SA (C. Au Peh); Sir Charles Gairdner Hospital, Nedlands, WA (A. Chakera); Royal North Shore Hospital, St Leonards (B. Cooper); Griffith University, Southport (J. Kurtkoti); Wesley Medical Research, Auchenflower (D. Langguth); Western Health, St. Albans Victoria (V. Levidiotis); Prince of Wales Hospital, Randwick NSW (G. Luxton); Austin Health, Heidelberg Victoria (P. Mount); Princess Alexandra Hospital, Woolloongabba, QLD (D. Mudge); Sunshine Coast University Hospital, Birtinya (E. Noble); Westmead Hospital, Westmead NSW (R. Phoon); Royal Brisbane and Women's Hospital, Herston QLD (D. Ranganathan); Concord Repatriation General Hospital, Concord (A. Ritchie); Monash Medical Centre, Clayton Victoria (J. Ryan); Liverpool Hospital, Liverpool, NSW (M. Suranyi).

**Austria**— National Coordinating Center: Medizinische Universitaet Graz, Graz (A. Rosenkranz); Landeskrankenhaus Feldkirch, Feldkirch (K. Lhotta); Medical University of Innsbruck, Innsbruck (A. Kronbichler).

**Belgium**— National Coordinating Center: Cliniques Universitaires Saint-Luc, Brussels (N. Demoulin); Centre Hospitalier Universitaire (CHU) de Liege, Liege (C. Bovy); Antwerp University Hospital (UZA), Edegem (R. Hellemans); Universite Libre de Bruxelles (ULB) - Hopital Erasme, Brussels (J. Hougardy); University Hospital (UZ) Leuven, Leuven (B. Sprangers); University Hospital Brussels, Brussels (K. Wissing).

**Canada**— National Coordinating Center: University of Toronto, Toronto (C. Pagnoux); St. Paul Hospital, Vancouver (S. Barbour); Centre de Recherche du Centre Hospitalier de l'Université de Montréal, Montreal (S. Brachemi); CISSS de la Monteregie-Centre – Hopital Charles LeMoyne, Greenfield Park (S. Cournoyer); University of Calgary, Calgary (L. Girard); Hospital Maisonneuve-Rosemont, Montreal (L. Laurin); Centre Hospitalier Universitaire de Sherbrooke, Sherbrooke (P. Liang); CHUQ-L'Hotel-Dieu de Quebec, Quebec City (D. Philibert); St. Josephs Healthcare, Hamilton (M. Walsh).

**Czech Republic**— Department of Nephrology, General University Hospital, Prague (V. Tesar); Rheumatology Institute, Prague (R. Becvar); University Hospital Olomouc, Olomouc (P. Horak); University Hospital Vinohrady, Prague (I. Rychlik).

**Denmark**— National Coordinating Center: Copenhagen University Hospital, Copenhagen (W. Szpirt); Odense University Hospital, Odense (H. Dieperink); Aalborg University Hospital, Aalborg (J. Gregersen); Aarhus University Hospital - Skejby, Aarhus (P. Ivarsen); Herlev Hospital, Herlev (E. Krarup); Sjaellands Universitetshospital Roskilde, Roskilde (C. Lyngsoe).

**France**— National Coordinating Center: CHU Bordeaux - Hospital Pellegrin, Bordeaux (C. Rigothier); CHU Angers, Angers (J. Augusto); CHU Lyon- Hopital Femme- Mere-Enfant, Bron (A. Belot); CHU de Toulouse - Hospital Rangueil, Toulouse (D. Chauveau); CHU de Brest - Hopital de la Cavale Blanche, Brest (D. Cornec); APHM - Hopital de la Conception, Marseille (N. Jourde-Chiche); CHU de Caen, Caen (M. Ficheux); Hopital Europeen Georges Pompidou, Paris (A. Karras); Hopitaux Civils de Colmar, Colmar (A. Klein); Hopitaux Prives de Metz, Metz (F. Maurier); Centre Hospitalier Boulogne sur Mer, Boulogne sur Mer (R. Mesbah); CHU Nimes – Hopital Caremeau, Nimes (O. Moranne); CHU Nantes Medicine Interne, Nantes (A. Neel); Centre Hospitalier de Valenciennes, Valenciennes (T. Quemeneur); Hospital Pitie Salpetriere, Paris (D. Saadoun); Hopital Cochin, Paris (B. Terrier); CHU de Grenoble, Grenoble Isere Cedex (P. Zaoui).

**Germany**— National Coordinating Center: University Clinic Heidelberg, Heidelberg (M. Schaier); University Clinic Mannheim, Mannheim (U. Benck); Clinic of Ludwigshafen am Rhein, Ludwigshafen (R. Bergner); University Clinic Jena, Jena (M. Busch); University Clinic Aachen, Aachen (J. Floege); University Clinic Cologne, Cologne (F. Grundmann); Medizinische Hochschule Hannover, Hannover (H. Haller); Klinikum Fulda, Fulda (M. Haubitz); Medius Clinic Kirchheim, Kirchheim-unter-Teck (B. Hellmich); University Hospital Tuebingen, Tuebingen (J. Henes); Nephrological Center Villingen-Schwenningen, Villingen-Schwenningen (B. Hohenstein); University Clinic Carl Gustav Carus, Dresden (C. Hugo); Klinikum Bad Bramstedt GmbH, Bad Bramstedt (C. Iking-Konert and F. Arndt); Asklepios Kinik, Hamburg (T. Kubacki and I. Kotter); University Clinic Schleswig-Holstein, Luebeck (P. Lamprecht); University Clinic Leipzig, Leipzig (T. Lindner and J. Halbritter); Charité - Universitaetsmedizin Berlin, Berlin (H. Mehling); Universität München – Großhadern, Munich (U. Schönermarck); University Clinic Freiburg, Freiburg (N. Venhoff); University Clinic Munich, Munich (V. Vielhauer); University

Clinic Essen, Essen (O. Witzke).

**Hungary**— Qualiclinic Kft, Budapest (I. Szombati); DEOEC Rheumatology Faculty, Debrecen (G. Szucs).

**Italy**— National Coordinating Center: IRCCS Azienda Ospedaliera Universitaria San Martino, Genova (G. Garibotto); ASST Santi Paolo e Carlo-Presidio Ospedale San Carlo, Milan (F. Alberici); Istituto Clinico Humanitas, Rozzano (E. Brunetta); IRCCS Ospedale San Raffaele, Milan (L. Dagna); Azienda Sanitaria Universitaria Integrata di Udine, Udine (S. De Vita); Azienda Ospedaliero-Universitaria Careggi, Florence (G. Emmi); AOU Ospedali Riuniti di Ancona, Torrette Ancona (A. Gabrielli); Azienda Ospedaliero Universitaria di Parma, Parma (L. Manenti); ASST di Monza-Ospedale San Gerardo, Monza (F. Pieruzzi); ASL Città di Torino - Ospedale San Giovanni Bosco, Torino (D. Roccatello); Azienda Unità Sanitaria Locale di Reggio Emilia, Reggio Emilia (C. Salvarani).**Japan**— National Coordinating Investigator: Prof. M. Harigai, Tokyo Women's Medical University, Tokyo; Kagawa University Hospital, Kagawa (H. Dobashi); Hokkaido University Hospital, Hokkaido (T. Atsumi); University of Miyazaki Hospital, Miyazaki (S. Fujimoto); Teikyo University Chiba Medical Center, Chiba (N. Hagino); National Hospital Organization Yokohama Medical Center, Yokohama (A. Ihata); Kyorin University Hospital, Tokyo (S. Kaname); Keio University Hospital, Tokyo (Y. Kaneko); Juntendo University Shizuoka Hospital, Shizuoka (A. Katagiri); Nagoya Medical Center, Aichi (M. Katayama); Yokohama City University Hospital, Kanagawa (Y. Kirino); National Hospital Organization Kanazawa Medical Center, Ishikawa (K. Kitagawa); Akita University Hospital, Akita City (A. Komatsuda); Teikyo University Hospital, Tokyo (H. Kono); Saitama Medical Center, Saitama (T. Kurasawa); National Hospital Organization Chiba East Hospital, Chiba (R. Matsumura); Saitama Medical University Hospital, Saitama (T. Mimura); Kobe University Hospital, Hyogo (A. Morinobu); Shimane University Hospital, Shimane (Y. Murakawa); Nagoya City University Hospital, Aichi (T. Naniwa); Toho University Omori Medical Center, Tokyo (T. Nanki); Hamamatsu University Hospital, Shizuoka (N. Ogawa); National Hospital Organization Tokyo Medical Center, Tokyo (H. Oshima); Okayama University Hospital, Okayama (K. Sada); Hiroshima University Hospital, Hiroshima (E. Sugiyama); Osaka Medical College Hospital, Osaka (T. Takeuchi); Toyama University Hospital, Toyama (H Taki); Juntendo University Hospital, Tokyo (N. Tamura); Tazuke Kofukai Medical Research Institute Kitano Hospital, Osaka (T. Tsukamoto); University of Tsukuba Hospital, Ibaraki (K. Yamagata); Okayama Saiseikai General Hospital, Okayama (M.

Yamamura).

**The Netherlands**— Erasmus MC, Rotterdam (P. van Daele); Groningen Universitair Medisch Centrum, Groningen (A. Rutgers); Leids Universitair Medisch Centrum, Leiden (Y. Teng).

**New Zealand**— National Coordinating Center: Dunedin Hospital, Dunedin (R. Walker); Christchurch Clinical Studies Trust, Christchurch (I. Chua); Auckland City Hospital, Auckland (M. Collins); Waikato Hospital, Hamilton (K. Rabindranath); North Shore Hospital, Takpuna, Auckland (J. de Zoysa).

**Norway**— National Coordinating Center: Akershus Universitetssykehus, Nordbyhagen (M. Svensson); Oslo Universitessykkehus, Oslo (B. Grevbo); University Hospital of North Norway, Tromso (S. Kalstad).

**Republic of Ireland**— National Coordinating Center: Beaumont Hospital, Dublin (M. Little); Cork University Hospital, Cork (M. Clarkson); St. Vincent's University Hospital, Dublin (E. Molloy).

**Spain**— Hospital Vall D Hebron, Barcelona (I. Agraz Pamplona); Hospital Sant Joan de Deu, Barcelona (J. Anton); Hospital Universitario Infanta Sofia, San Sebastian de los Reyes, Madrid (V. Barrio Lucia); Hospital Da Costa, Burela (S. Ciggaran); Hospital Clinic Barcelona – Autoimmune Diseases Department, Barcelona (M. Cinta Cid); Fundacio Puigvert, Barcelona (M. Diaz Encarnacion); Hospital Universitari de Bellvitge, Barcelona (X. Fulladosa Oliveras); Hospital del Mar, Barcelona (M. Jose Soler); Hospital Germans Trias i Pujol, Badalona (H. Marco Rusinol); Hospital 12 de Octubre, Madrid (M. Praga); Hospital Clinic Barcelona, Barcelona (L. Quintana Porras); Hospital Universitari Arnau de Vilanova, Lleida (A. Segarra).

**Sweden**— National Coordinating Center: Karolinska University Hospital, Stockholm (A. Bruchfeld); Linköping University, Linköping (M. Segelmark); Uppsala University Hospital, Uppsala (I. Soveri); Örebro University Hospital, Örebro (E. Thomaidi); Skane University Hospital, Malmo (K. Westman).

**Switzerland**— National Coordinating Center: Kantonsspital St. Gallen, St. Gallen (T. Neumann); CHUV Lausanne, Lausanne (M. Burnier); University Hospital Basel, Basel (T. Daikeler); Hôpital Fribourgeois, Fribourg (J. Dudler); Immunologie- Zentrum Zürich, Zürich (T. Hauser); Universitätsspital Zürich, Zürich (H. Seeger); Inselspital, Universitätsspital Bern, Bern (B. Vogt).

**United Kingdom**— National Coordinating Center: Addenbrooke's Hospital - Cambridge University Hospitals, Cambridge (D. Jayne); Leicester General Hospital, Leicester (J. Burton and R. Al Jayyousi); Leeds Childrens Hospital, Leeds (T. Amin); Leeds Teaching Hospitals NHS Trust, Leeds (J. Andrews); Freeman Hospital, Newcastle upon Tyne (L. Baines); Great Ormond Street Hospital for Children, London (P. Brogan); Southend University Hospital, Westcliff on Sea (B. Dasgupta); Kent and Canterbury Hospital, Canterbury – Kent (T. Doulton); Royal Berkshire Hospital, Reading, Berkshire (O. Flossmann); University Hospital of Wales, Cardiff (S. Griffin); Royal Liverpool University Hospital, Liverpool (J. Harper); University of Birmingham, Birmingham (L. Harper); University Aberdeen, Aberdeen (D. Kidder); Russells Hall Hospital, Dudley (R. Klocke); Queens Medical Centre, Nottingham (P. Lanyon); Nuffield Orthopaedic Centre, Oxford (R. Luqmani); Whytemans Brae Hospital, Fife (J. McLaren); St Helier Hospital, Carshalton (D. Makanjuola); Alder Hey Children's NHS Foundation Trust, Liverpool (L. McCann); Basildon University Hospital, Basildon (A. Nandagudi and S. Selvan); Salford Royal NHS Foundation Trust Manchester, Salford (E. O'Riordan); University of Manchester, Manchester Royal Infirmary, Manchester (M. Patel); Queen Elizabeth University Hospital, Glasgow (R. Patel); Imperial College Healthcare NHS Trust, London (C. Pusey); The Royal London Hospital, London (R. Rajakariar); Bristol Royal Infirmary, Bristol (J. Robson); Guy's and St Thomas's NHS Foundation Trust, London (M. Robson); UCL Centre for Nephrology Royal Free, London (A. Salama); Royal Devon and Exeter Hospital, Exeter (L. Smyth); Raigmore Hospital, Inverness (J. Sznajd); Dorset County Hospital, Dorchester (J. Taylor).

**United States of America**—University of Pennsylvania, Philadelphia (P. Merkel and A. Sreih); Winthrop University Hospital, Mineola (E. Belilos); Columbia University Medical Center, New York (A. Bomback); Virginia Mason Medical Center, Seattle (J. Carlin); University of South Florida, Tampa (Y. Chang Chen Lin); University of North Carolina Hospitals, Chapel Hill (V. Derebail); MedStar Georgetown University Hospital, Washington (S. Dragoi); University of Chicago Medical Center Rheumatology, Chicago (A. Dua); Cedars-Sinai Medical Center, Los Angeles (L. Forbess); Johns Hopkins Bayview Medical Center, Baltimore (D. Geetha); University of Michigan, Ann Arbor (P. Gipson); Rhode Island Hospital, Providence (R. Gohh); Brookview Hills Research Associates, Winston-Salem (G. T. Greenwood); Indiana University Nephrology, Indianapolis (S. Hugenberg); Western Washington Arthritis Clinic, Bothell (R. Jimenez); Northwest Louisiana Nephrology, Shreveport (M. Kaskas); University of California, Los Angeles,

Santa Monica (T. Kermani); Altoona Center for Clinical Research, Duncansville (A. Kivitz); University of Utah, Salt Lake City (C. Koening); Cleveland Clinic, Cleveland (C. Langford); Northwell Health, Great Neck (G. Marder); University of Kentucky Medical Center, Lexington (A. Mohamed); Boston University, Boston (P. Monach); Arizona Kidney Disease and Hypertension Center Flagstaff, Flagstaff (N. Neyra); Articularis Healthcare Group, Charleston (G. Niemer); Massachusetts General Hospital, Boston (J. Niles); East Carolina University, Greenville (R. Obi); Renal Disease Research Institute, Dallas (C. Owens); Washington University School of Medicine, St. Louis (D. Parks); Colorado Kidney Care, Denver (A. Podoll); Ohio State University, Columbus (B. Rovin); San Francisco General Hospital Dialysis Center, San Francisco (R. Sam); Rheumatology Associates of North Alabama, Huntsville (W. Shergy); Boise Kidney & Hypertension, PLLC – Meridian, Caldwell (A. Silva); Mayo Clinic - Division of Pulmonary & Critical Care Medicine, Rochester (U. Specks); Hospital for Special Surgery, New York (R. Spiera); University of Kansas Medical Center, Kansas City (J. Springer); University of Colorado Denver - School of Medicine, Aurora (C. Striebich); Arizona Arthritis & Rheumatology Research, Phoenix (A. Swarup); University of Minnesota, Minneapolis (S. Thakar); Emory University School of Medicine, Atlanta (A. Tiliakos); Arthritis, Autoimmune and Allergy LLC, Daytona Beach (Y. Tsai); University of Texas Health Sciences Center, Houston (D. Waguespack); Allegheny General Hospital, Pittsburgh (M. Chester Wasko).

#### 2. ELIGIBILITY CRITERIA

The screening period for study eligibility was not to exceed 14 days. Eligible patients were at least 12 years old and had newly-diagnosed or relapsing granulomatosis with polyangiitis or microscopic polyangiitis, according to the Chapel Hill Consensus Conference definitions,[1] for whom treatment with cyclophosphamide or rituximab was indicated, tested positive for antibodies to either proteinase-3 (PR3) or myeloperoxidase (MPO), had an estimated glomerular filtration rate (eGFR) of at least 15 mL/min/1.73 $m^2$, had at least one major or three non-major items, or at least two renal items of hematuria and proteinuria on the Birmingham Vasculitis Activity Score (BVAS). The BVAS includes disease activity assessment in 9 organ systems, it ranges from 0 to 63, with higher scores indicating more organ involvement.[2]

Patients were excluded if they had alveolar hemorrhage requiring invasive pulmonary ventilation anticipated to last beyond screening, any other multisystem autoimmune disease, a coagulopathy or bleeding disorder, required dialysis or plasma exchange within 12 weeks prior to screening, had a kidney transplant, or had received any of the following: cyclophosphamide within 12 weeks prior to screening, rituximab within 12 months prior to screening (or 6 months with B-cell reconstitution, CD19 count $>0.01x10^9$/L), a cumulative dose of intravenous glucocorticoids greater than 3 g within 4 weeks, or oral glucocorticoids of more than 10 mg per day prednisone (or equivalent) for more than 6 weeks continuously prior to screening.

#### 3. STUDY TREATMENTS

Patients in both treatment groups received either intravenous or oral cyclophosphamide or intravenous rituximab as follows: intravenous cyclophosphamide at a dose of 15 mg/kg up to a maximum of 1.2 g on day 1 and at weeks 2, 4, 7, 10, and 13; oral cyclophosphamide at a dose of 2 mg/kg up to a maximum of 200 mg per day for 14 weeks (Supplemental Table S1); or intravenous rituximab at a dose of 375 mg/m2 per week for 4 weeks. From week 15 onward, cyclophosphamide was followed by oral azathioprine at a target dose of 2 mg/kg per day. If azathioprine was not tolerated, mycophenolate mofetil at a target dose of 2 g/day may have been given. If mycophenolate mofetil was not tolerated or not available, enteric coated mycophenolate sodium may have been given at a target dose of 1440 mg/day.

### 4. STUDY ASSESSMENTS

#### 4.1. Birmingham Vasculitis Activity Score (BVAS)

BVAS v3 employs 52 items of vasculitis activity grouped into 10 organ systems (shown below) with each item defined and given a weighted score with a maximum score for each organ group, and a maximum total score of 63. If all scored items were present but not worse than the preceding assessment a persistent box is scored with a lower weighted score.[2] Three modifications to BVAS v3 were made for the ADVOCATE trial, all items were categorized as either major or minor and an additional box (red cell casts of glomerulonephritis) was added as a major item to simplify the categorization of kidney vasculitis and to align with the version of BVAS, BVAS/WG used for the pivotal RAVE trial that supported approval of rituximab for GPA/MPA.[3] The persistent box was removed because, in the opinion of the investigators, the presence or absence of an item of persisting disease activity, that was not worse, was associated with higher inter-observer variation than whether an item was new or worse.

The BVAS was completed during screening, and at weeks 4, 10, 16, 26, 39, 52, and 60. As part of the 2026 readjudication, the BVAS scores at weeks 26, 39, and 52 and at any unscheduled or early termination visits occurring between weeks 26 and 52 were re-evaluated.

The BVAS organ systems are shown below. Major items are indicated in bold italics. For study eligibility, a patient had to have at least one major item, at least 3 minor items, or at least the two renal items of hematuria and proteinuria. For the week 4 BVAS assessment, disease activity within 7 days prior to the week 4 visit was assessed. This was done because the typical 28 days would potentially include the baseline assessment. For all other visits, disease activity within 28 days prior to the visit was assessed. Relapses were assessed at all study visits after week 4. As part of the 2026 readjudication, relapse was reassessed between weeks 26 and 52.

Body Systems:

1. General
   - Myalgia
   - Arthralgia / arthritis
   - Fever ≥38 °C
   - Weight loss ≥2 kg
2. Cutaneous
   - Infarct
   - Purpura
   - Ulcer
   - ***Gangrene***
   - Other skin vasculitis
3. Mucous membranes / eyes
   - Mouth ulcers
   - Genital ulcers
   - Adnexal inflammation
   - Significant proptosis
   - ***Scleritis / Episcleritis***
   - Conjunctivitis / Blepharitis / Keratitis
   - Blurred vision
   - Sudden visual loss
   - Uveitis
   - ***Retinal changes (vasculitis / thrombosis / exudate / haemorrhage)***
4. Ear Nose & Throat
   - Bloody nasal discharge / crusts / ulcers / granulomata
   - Paranasal sinus involvement
   - Subglottic stenosis
   - Conductive hearing loss
   - ***Sensorineural hearing loss***
5. Chest
   - Wheeze
   - Nodules or cavities
   - Pleural effusion / pleurisy
   - Infiltrate
   - Endobronchial involvement
   - ***Massive hemoptysis / alveolar hemorrhage***
   - ***Respiratory failure***

6. Cardiovascular
    - Loss of pulses
    - Valvular heart disease
    - Pericarditis
    - Ischemic cardiac pain
    - Cardiomyopathy
    - Congestive cardiac failure
7. Abdominal
    - Peritonitis
    - Bloody diarrhea
    - ***Ischemic abdominal pain***
8. Renal
    - Hypertension
    - Proteinuria >1+ or >0.2 g/g creatinine
    - Hematuria ≥10 RBCs/hpf
    - Serum creatinine 125-249 µmol/L
    - Serum creatinine 250-499 µmol/L
    - Serum creatinine ≥500 µmol/L
    - ***Rise in serum creatinine >30% or fall in creatinine clearance >25%***
9. Nervous system
    - Headache
    - ***Meningitis***
    - Seizures (not hypertensive)
    - ***Cerebrovascular accident***
    - Organic confusion
    - ***Spinal cord lesion***
    - ***Cranial nerve palsy***
    - ***Sensory peripheral neuropathy***
    - ***Mononeuritis multiplex***
10. Other
    - ***RBC casts and/or glomerulonephritis***

#### 4.2. Glucocorticoid Toxicity Index

Glucocorticoid toxicity was analyzed using the Glucocorticoid Toxicity Index (GTI) version 2.0,[4] an instrument upgrade from the original GTI version 1.0.[5] Data for completion of the GTI were collected on day 1 (pre-dosing) and at weeks 13 and 26. The GTI 2.0 instrument provides two GTI scores: the Cumulative Worsening Score (CWS) and the Aggregate Improvement Score (AIS).

The GTI-CWS captures cumulative glucocorticoid toxicity regardless of whether it is permanent or transient. The GTI-CWS can only increase or remain the same over time. A lower score indicates lower glucocorticoid toxicity.

The GTI-AIS captures both worsening and improvement in glucocorticoid toxicity. New or worsening toxicities contribute a positive score and improvement in existing toxicities contributes a negative score. A lower score indicates lower glucocorticoid toxicity.

The GTI components and scoring are provided in Table S3. Definitions of individual items are provided in the original publication.[5]

#### 4.3. Other Study Assessments

##### 4.3.1. Health-Related Quality of Life

Health-related Quality of Life was assessed with the Medical Outcomes Study Short Form 36 version 2 survey[6] and the EQ-5D-5L questionnaire.[7] These assessments were completed on day 1 (pre-dosing), and weeks 4, 10, 16, 26, 39, 52, and 60.

##### 4.3.2. Renal Function

Serum creatinine, for estimated glomerular filtration rate (eGFR) calculation, was measured on Day 1, and weeks 1, 2, 3, 4, 7, 10, 13, 16, 20, 26, 32, 39, 45, 52, and 60 over the study course. eGFR (according to the Modification of Diet in Renal Disease equation[8]) in mL/min/1.73 $m^2$ = $175\times(\text{serum creatinine, mg/dL})^{-1.154}\times(\text{age, years})^{-0.203}\times(0.742 \text{ if female})\times(1.212 \text{ if black})$. For Japanese patients, the following equation was used: eGFR (mL/min/1.73 $m^2$) = $194\times(\text{serum creatinine, mg/dL})^{-1.094}\times(\text{age, years})^{-0.287}\times(0.739 \text{ if female})$.[9] The Modified Schwartz equation[10] was used for adolescents: eGFR = (0.413 x Height [in cm]) / serum creatinine (in mg/dL).

First morning urine samples for albumin, monocyte chemoattractant protein-1, and creatinine measurements were collected on day 1 (pre-dosing) and weeks 1, 2, 4, 13, 26, 39, 52, and 60. Urine albumin was measured by a nephelometric assay, monocyte chemoattractant protein-1 by ELISA, and creatinine by a kinetic colorimetric assay in a central laboratory (Medpace Reference Laboratory in Cincinnati, OH, USA; Leuven, Belgium; and Singapore). The vasculitis damage index was completed at screening and weeks 26, 52, and 60.[11]

###### 4.3.3. Safety

Safety was monitored at each study visit by assessing adverse events and laboratory data. The prospectively identified search terms for the glucocorticoid-related adverse event analysis are provided in Table S12.

###### 4.3.4. Compliance with Study Medication

Patient compliance with taking study medication was assessed based on returned capsule counts at each study visit.

#### 5. CRITERIA FOR ADJUSTING CYCLOPHOSPHAMIDE DOSE

The cyclophosphamide dose was determined by four factors: patient age, eGFR, white blood cell (WBC) count at the study visit, and WBC count nadir in between dose pulses (where applicable). The dose for oral and IV cyclophosphamide, based on age and eGFR, is provided in **Table S1**.

For those receiving IV cyclophosphamide, the dose was to be reduced further based on the WBC count as follows:

- WBC count assessed just prior to the IV dose:
  - If ≥ 3.5 x $10^9$/L, dosing according to **Table S1** was to be given;
  - If < 3.5 x $10^9$/L, the dose was to be postponed until the WBC count was ≥ 3.5 x $10^9$/L and then the dose from **Table S1** was to be reduced by another 25%;
- WBC count nadir in between IV cyclophosphamide doses:
  - If > 3 x $10^9$/L, dosing according to **Table S1** was to be given;
  - If 2 to 3 x $10^9$/L, the dose from **Table S1** was to be reduced by 20%;
  - If 1 to 1.9 x $10^9$/L, the dose from **Table S1** was to be reduced by 40%;
  - If < 1 x $10^9$/L, the next dose was to be withheld and further dosing was only to be given if the WBC was > 3 x $10^9$/L.

For those receiving oral cyclophosphamide, the dose should be reduced further based on the WBC count as follows:

- If WBC count was < 3.5 x $10^9$/L, dosing was to be withheld. When WBC count returned to ≥ 3.5 x $10^9$/L for two consecutive tests, or ≥ 5 x $10^9$/L on a single test, oral cyclophosphamide was to be restarted at 25 mg less than the dose from Table S1**.** WBC count was to be monitored weekly after this episode. If only 50 mg dose units were available, oral cyclophosphamide was to be restarted at 50 mg less than dose from **Table S1**.
- If WBC count was < 1 x $10^9$/L, or < 3.5 x $10^9$/L for more than 2 weeks, dosing was to be withheld. When WBC count returned to ≥ 3.5 x $10^9$/L for two consecutive tests or ≥ 5 x $10^9$/L on a single test, oral cyclophosphamide was to be restarted at 50 mg less than the dose from **Table S1**. The WBC count was to be monitored weekly after this episode.

Consideration was to be given for granulocyte colony stimulating factor administration, fungal prophylaxis, or other precautions.

- If WBC count decreased markedly without overt leukopenia, e.g., WBC count < 6 x $10^9$/L and at least 2 x $10^9$/L lower than before, weekly WBC count monitoring should have been done and the oral cyclophosphamide dose from **Table S1** reduced by 25 mg (or 50 mg if 25 mg dose units were not available) if the WBC count continued to decrease.

The last IV cyclophosphamide dose was to be given at the week 13 visit. The last oral cyclophosphamide dose was to be given the day prior to week 15.

#### 6. CRITERIA FOR GLUCOCORTICOID USE

The study protocol envisioned the use of some glucocorticoids in both groups as a function of administration during screening and prior to randomization; as co-administration with rituximab (to prevent hypersensitivity reactions per the rituximab prescribing information), and to manage non-vasculitis reasons (as detailed below). Such glucocorticoid use was balanced between the two groups (see **Table S9 and Table S10**). To the extent that glucocorticoids were allowed under certain conditions, it was specified in the protocol that such use should be limited.

##### 6.1. Prior to the Screening Period

Patients with severe ANCA-associated vasculitis were allowed per protocol to have received intravenous (IV) glucocorticoids at a cumulative dose up to 3 g methylprednisolone equivalent in the 4-week period prior to screening. Patients were also allowed to have received oral glucocorticoids in the 6-week period prior to screening. However, patients were not eligible for screening if they had received treatment with moderate or high dose glucocorticoids continuously for more than 6 weeks; moderate to high dose was defined as greater than 10 mg prednisone-equivalent per day.

##### 6.2. During the Screening Period

During the screening period of the study (which was not to exceed 14 days), IV glucocorticoids were allowed for patients with severe vasculitis, as long as the cumulative dose for the 4-week period prior to screening plus the IV dose(s) given during the screening period did not exceed 3 g methylprednisolone equivalent. During the screening period of the study, oral glucocorticoids were allowed for patients with severe vasculitis. If a patient received oral glucocorticoids during the screening period, the dose needed to be tapered to a dose that did not exceed 20 mg prednisone equivalent on Day 1 (first dosing day) of the study.

##### 6.3. During the Treatment Period

Glucocorticoid treatment was allowed as pre-medication for the rituximab infusions, typically 100 mg methylprednisolone equivalent prior to each infusion. The majority of patients who received IV glucocorticoids during the first 4 weeks in **Table S10** received it as pre-medication. Only 19 patients in the prednisone group and 21 in the avacopan group received IV glucocorticoids that were not for pre-medication.

If a patient was still taking a dose of ≤20 mg prednisone-equivalent oral glucocorticoids (other than prednisone study medication) on Day 1 of the study, the dose of the oral glucocorticoids had to be tapered to zero over a 4-week period after Day 1. The protocol allowed glucocorticoid use for non-vasculitis reasons, e.g., replacement glucocorticoids for adrenal insufficiency, treatment of allergic reaction, treatment of gout. Patients who experienced a relapse of their vasculitis during the study could be treated with IV glucocorticoids (typically 0.5 to 1 g methylprednisolone per day for 3 days) and/or oral glucocorticoids, tapered according to the patient's condition. These patients could continue study drug treatment and could continue in the study.

Patients who experienced worsening of disease during the study that involved a major item in the BVAS could be treated with IV glucocorticoids (typically 0.5 to 1 g methylprednisolone per day for 3 days) and/or oral glucocorticoids, tapered according to the patient's condition. Worsening not involving a major item in the BVAS could be treated with a short burst (i.e., not more than 2 weeks) of oral glucocorticoids, at a maximum dose of 20 mg prednisone equivalent. Patients experiencing worsening of disease could continue study drug treatment and could continue in the study.

Patients who had one or more major items in the BVAS before study entry, and who did not show an improvement or stabilization of these major items within the first 4 weeks of the study, could receive additional IV or oral glucocorticoids, tapered according to the patient’s condition.

#### 7. 2026 READJUDICATION COMMITTEE

To address concerns raised regarding the process used to adjudicate the primary outcomes of 9 participants in the 2019 analysis, a fully blinded, independent, readjudication (the 2026 readjudication) of the primary endpoints in the ADVOCATE trial was performed by the Duke Clinical Research Institute (DCRI) Clinical Events Classification (CEC) group. The 2026 readjudication committee was instructed to adjudicate cases in accordance with an Adjudication Charter that followed processes and assessment parameters similar to those of the original ADVOCATE Adjudication Committee Charter.

In contrast to the original adjudication committee, the 2026 adjudicators were asked to adjudicate week 26, week 39, and week 52 assessments and relapse events, as well as any unscheduled or early termination visits that occurred between weeks 26 and 52. They could not issue queries to trial sites, although they had access to queries issued by the original adjudication committee. The 2026 committee was trained through the case-based program offered by Professor Raashid Luqmani at Oxford University; the original committee was trained by Dr. David Jayne.

The original adjudication committee resolved reviewer disagreements verbally during meetings, without documenting those discussions in meeting minutes. Although duplicate versions of the original adjudication forms were used for the 2026 readjudication, the DCRI process required documentation of all adjudication decisions related to the BVAS, VDI, and relapse. Discrepancies between adjudicators in the 2026 readjudication were referred to resolution meetings, and commentary documenting the final decision was added to the adjudication record. In addition, Dr. Luqmani performed quality control review of 10% of DCRI-adjudicated cases; a similar process was not used in the original adjudication.

Because the 2026 readjudication occurred after trial completion, Medpace, the Contract Research Organization involved in ADVOCATE, prepared new datasets for DCRI that removed prior adjudication assessments.

### 8. STATISTICAL ASPECTS

The two primary efficacy end points were the proportion of patients who (1) achieved remission at week 26 and (2) sustained remission at week 52. For each primary efficacy end point, non-inferiority of the avacopan group to the prednisone group was tested using a one-sided testing procedure (or equivalently, using a one-sided confidence interval [CI]) and a non-inferiority limit of –0.20. Subject to the procedures for controlling the overall Type I error, a test for avacopan superiority could also be performed for each primary efficacy end point. To preserve the overall Type I error at the 0.05 level for testing two primary efficacy end points and two tests of non-inferiority and superiority for each primary efficacy end point, the hierarchical procedure was implemented: (1) non-inferiority at week 26, (2) non-inferiority at week 52, (3) superiority at week 52, and (4) superiority at week 26. The Summary score test[12] was used for both non-inferiority and superiority tests of the stratified analyses of the two primary efficacy end points. The stratification factors were the same factors used in the randomization stratification: either newly diagnosed or relapsing vasculitis, PR3 or MPO-ANCA, and cyclophosphamide or rituximab treatment. Summary score estimates[12] of the common difference in remission rates between the avacopan and prednisone groups were provided using inverse-variance stratum weights and Miettinen-Nurminen (score) confidence limits for the common difference in remission rates. Primary end point analyses were conducted after all patients have completed the 52-week treatment period. No interim analyses were performed.

The primary efficacy analyses were conducted on the modified intention-to-treat population defined as all randomized patients who received at least one dose of study medication (avacopan/placebo). Per-protocol analyses were also conducted where patients with major eligibility criteria deviations and missing week 26 data were excluded, and non-remission was imputed for patients who were <75% compliant with taking study medication based on returned capsule count, or used non-allowed immunosuppressants during the treatment period.

For the primary end points, if the lower bound of the 2-sided 95% confidence interval for the difference (avacopan group minus prednisone group) in remission rate was greater than –0.20, the avacopan group would be considered not inferior to the prednisone group. If the lower bound was greater than 0.0, the avacopan group would be considered superior to the prednisone group. For the primary end points, missing data at week 26 and week 52 were imputed as not being in remission.

For the week 26 remission end point, 10 patients in each group had missing data. For the week 52 sustained remission end point, 15 patients in the avacopan group and 12 patients in the prednisone group had missing data.

Other binary end points were analyzed similarly to the primary end points. Continuous variables were analyzed using mixed effects models for repeated measures (MMRM). The least squares means (LSMs), standard errors, and CIs are from models incorporating treatment group, visit, treatment-by-visit interaction and stratification factors as covariates. Longitudinal measurements from the same patients were considered as repeated measure units in the model. In the MMRM model, missing data were not imputed. This analysis is unbiased under the missing at random assumption. A compound symmetry covariance matrix was used to model the within-subject variance-covariance structure for the model errors. For the secondary end points, point estimates and 95% CIs are presented as no multiple comparison adjustments were made and no definitive conclusions can be drawn.

### 9. DATA MONITORING COMMITTEE

An independent external Data Monitoring Committee oversaw the study and reviewed safety data over the course of the study. The Data Monitoring Committee recommended additional monitoring for potential hepatotoxicity and WBC count decreases. These recommendations were added by protocol amendment. At all its meetings, the Data Monitoring Committee recommended continuation of the study.

### 10. FIGURES

**Figure S1. Subgroup Analysis of Remission at Week 26 (A) and Sustained Remission at Week 52 (B) – 2026 Adjudication**

A

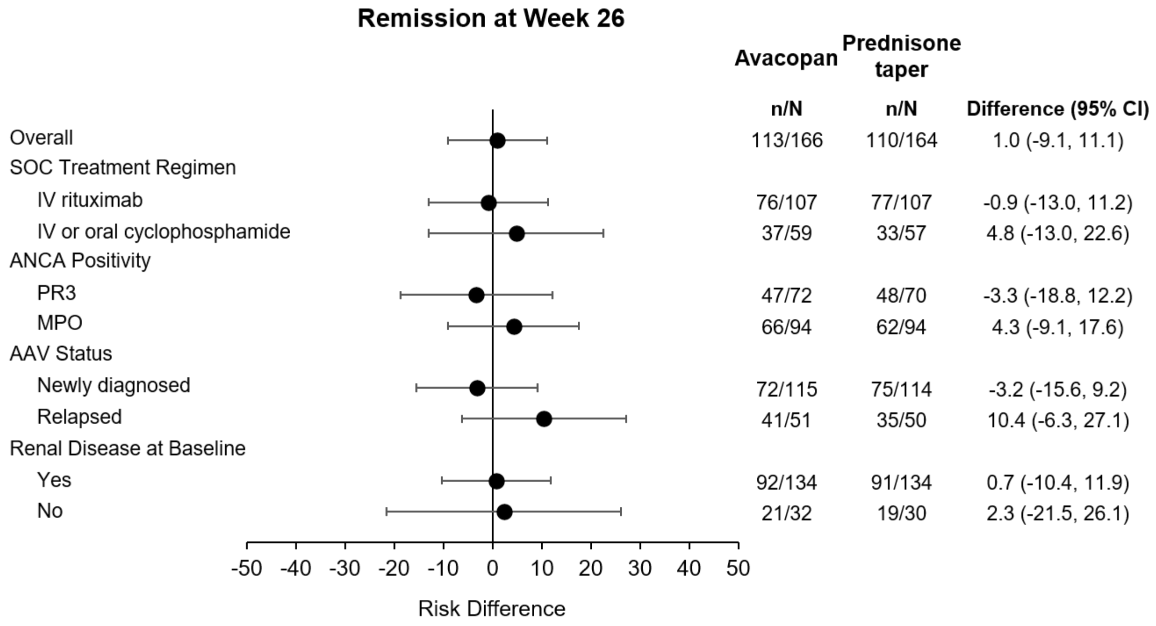

B

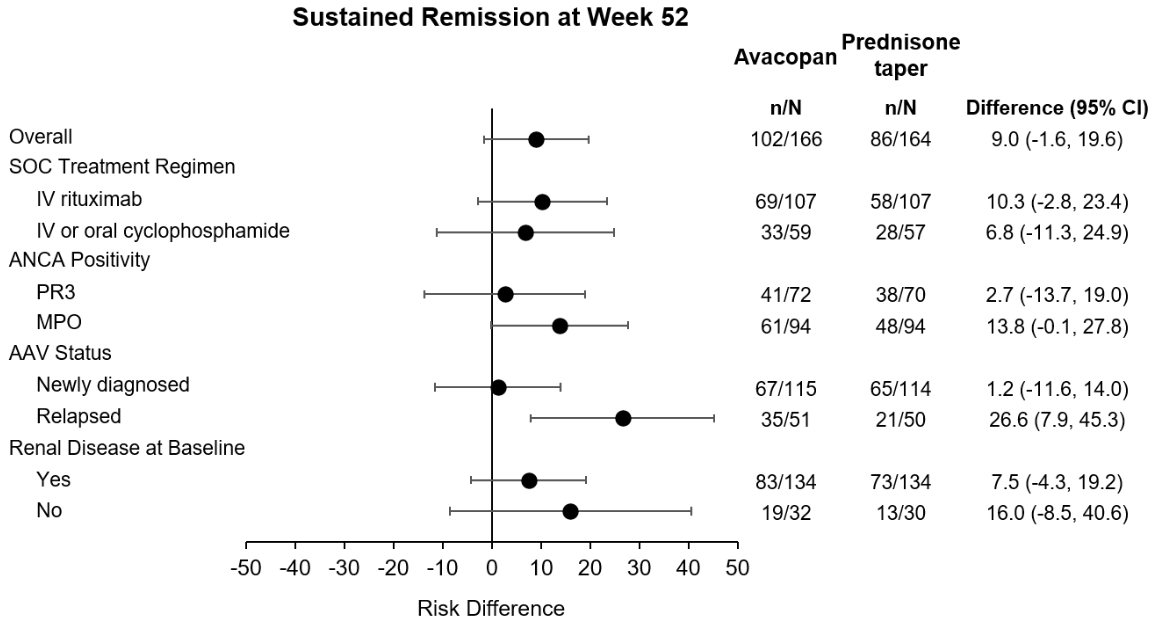

AAV, ANCA-associated vasculitis; ANCA, anti-neutrophil cytoplasmic antibody; CI, confidence interval; IV, intravenous; MPO, myeloperoxidase; PR3, proteinase 3; SOC, standard of care.

**Figure S2.** **Kaplan-Meyer Plot of Time to Relapse – 2019 Adjudication**

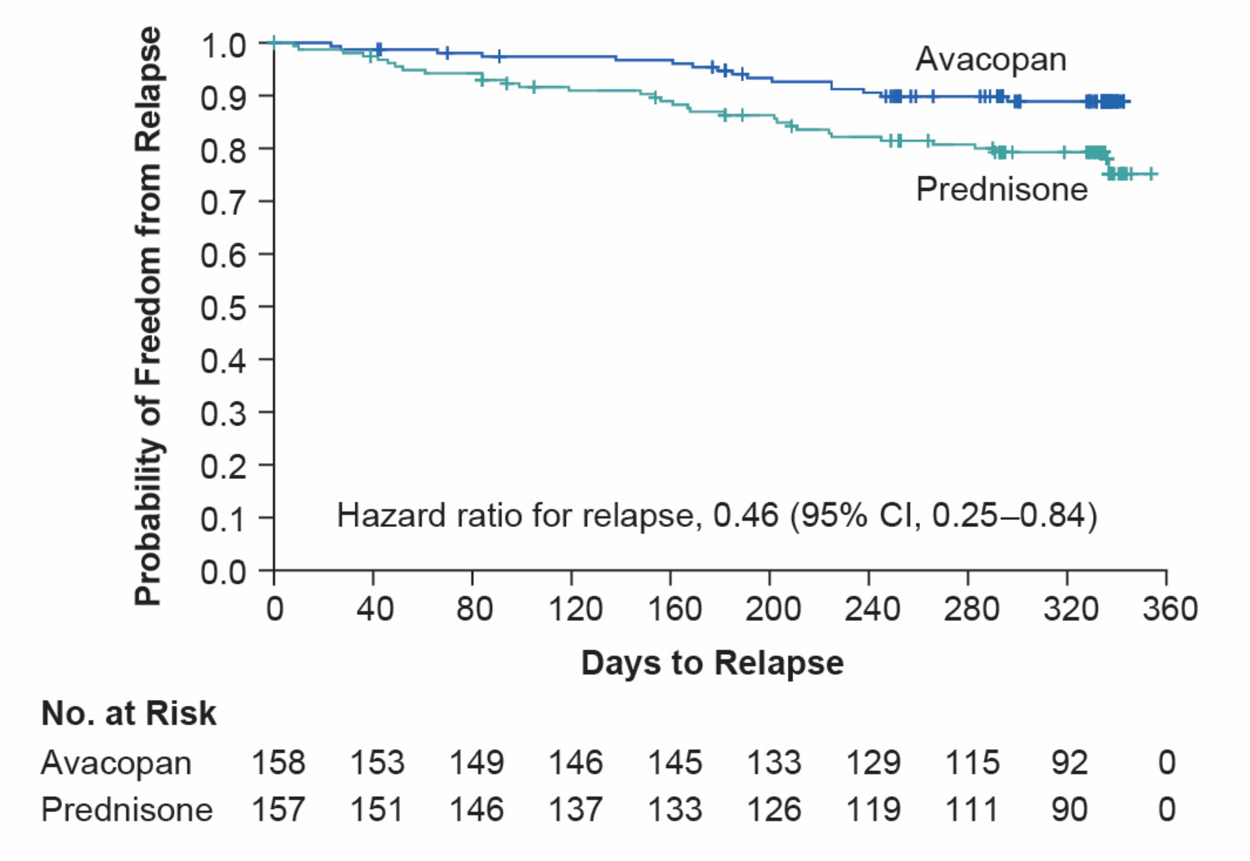

The hazard ratio for relapse after previous achievement of a BVAS=0 at any time point (avacopan vs. prednisone) was 0.46 (95% CI, 0.25, 0.84) according to the 2019 adjudication. As the BVAS for each participant was only assessed between weeks 26 and 52 in the 2026 readjudication, this analysis of relapse risk after achieving a BVAS=0 at any timepoint in the trial was not re-evaluated following the 2026 readjudication; instead, relapse after achieving remission at week 26 was assessed in the 2026 readjudication, as it was in the 2019 adjudication.

Figure reused from retracted article.[13,14]

Relapse was defined as worsening of disease after previous achievement of BVAS=0 involving: 1 or 2 minor BVAS items recorded at 2 consecutive study visits; 1 or more major BVAS items; or 3 or more minor BVAS items. A total of 16 of 158 patients (10.1%) in the avacopan group and 33 of 157 patients (21.0%) in the prednisone group had relapses. A test of proportionality was performed by incorporating a time-varying covariate in the Cox regression model by creating an interaction of the treatment groups and log of the time to relapse. The Wald chi-square test for the interaction term was 0.48. The corresponding *P* value was 0.49, which indicates no significant evidence of nonproportionality of the hazard. Tick marks indicate censored data.

**Figure S3. Estimated Glomerular Filtration Rate**

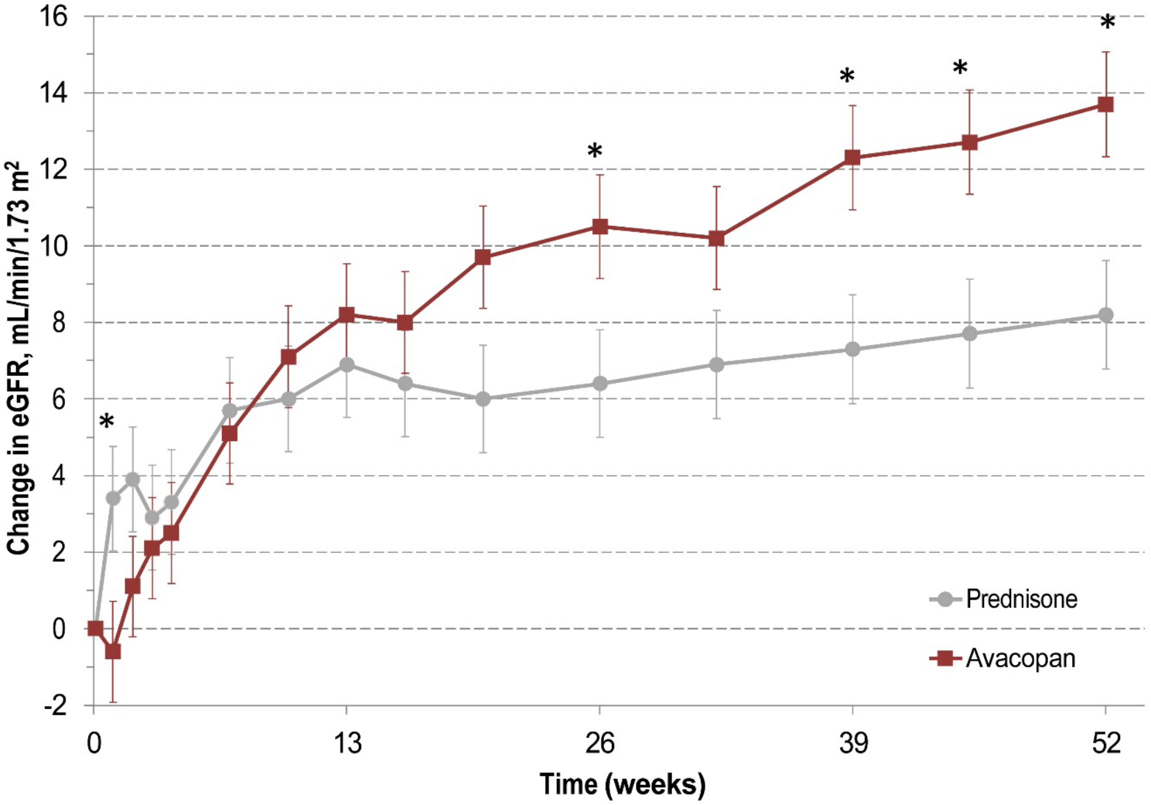

Change from baseline over the course of the 52-week treatment period in eGFR in patients with renal disease and eGFR <30 mL/min/1.73 $m^2$ at baseline; the mean ± standard error of mean baseline eGFR was 21.6 ± 0.66 and 21.1 ± 0.58 mL/min/1.73 $m^2$ in the prednisone and avacopan groups, respectively.

Data are presented as least squares means ± standard error of the means.

*Indicates where 95% confidence intervals of the differences between the avacopan and prednisone group do not include 0. The two treatment groups were compared by mixed effects model for repeated measures analysis with treatment group, study visit, treatment-by-visit interaction, and randomization stratum as factors and baseline as covariate.

**Figure S4. Mean Daily Total Oral Prednisone-Equivalent Glucocorticoid Dose (in mg) by Study Week by Treatment Group**

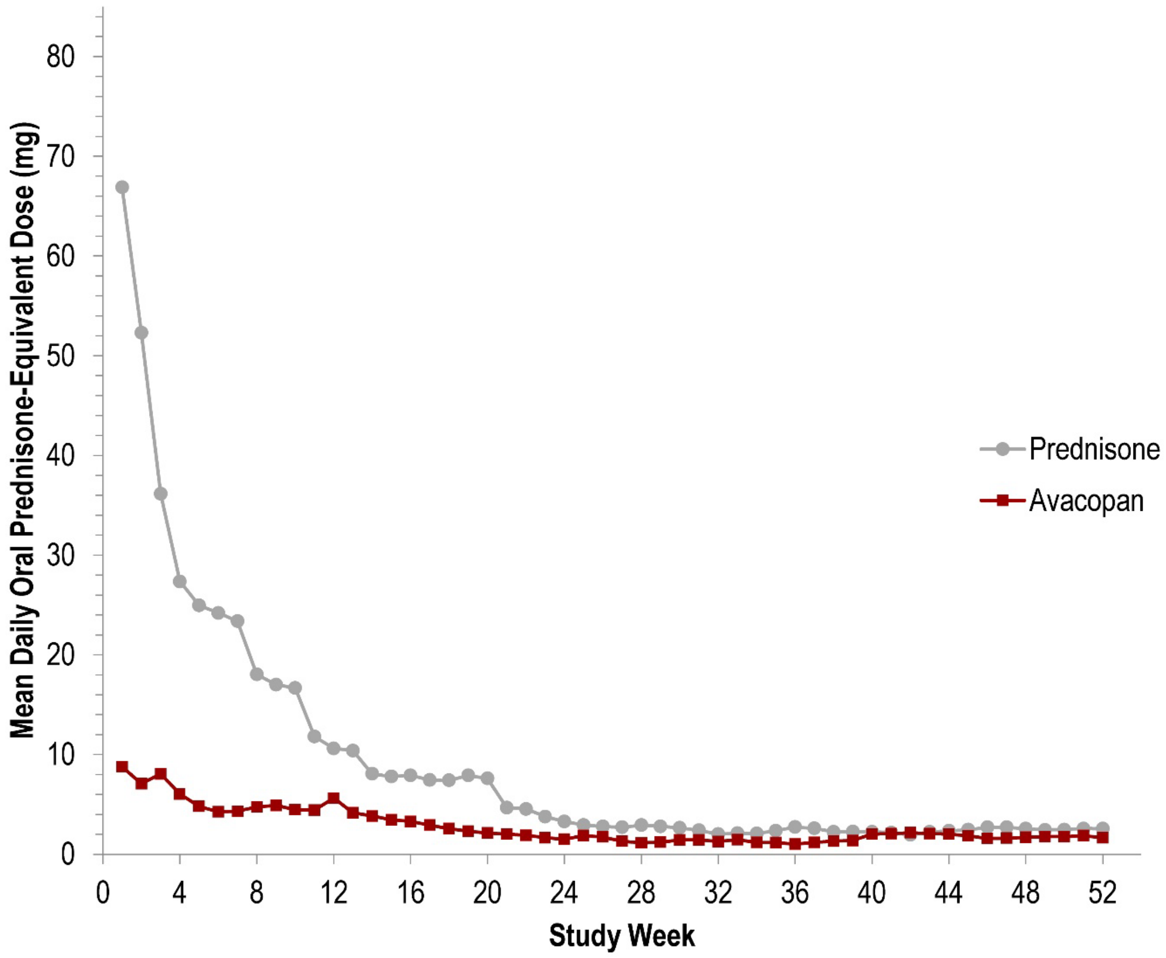

The weekly mean values were calculated based on all recorded systemic oral glucocorticoid use by all patients in the respective treatment group at the start of each study week.

**Figure S5. Mean Daily Total Prednisone-Equivalent Glucocorticoid Dose (in mg) by Study Week by Treatment Group**

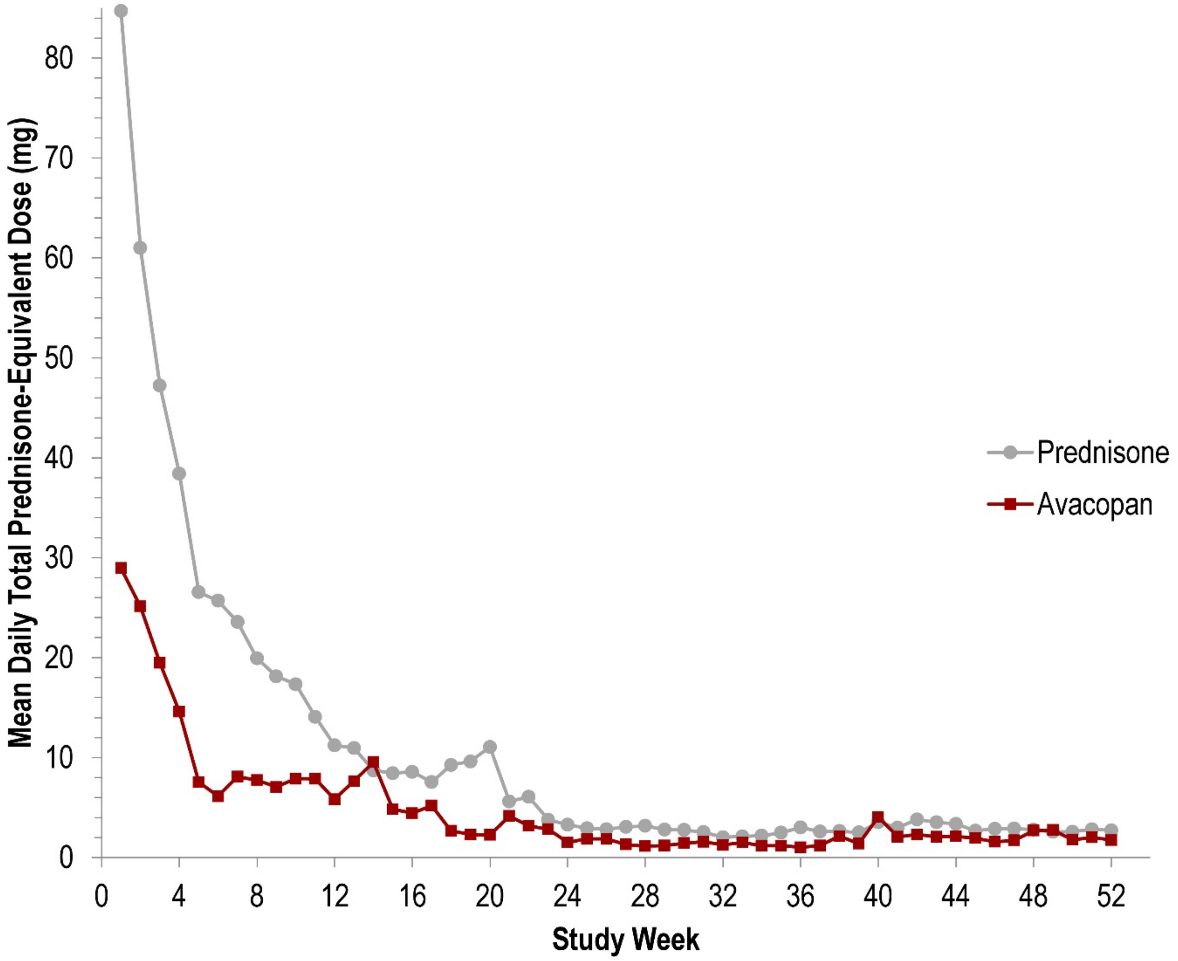

The weekly mean values were calculated based on all recorded systemic (oral or intravenous) glucocorticoid use by all patients in the respective treatment group at the start of each study week.

**Figure S6. Short Form 36**

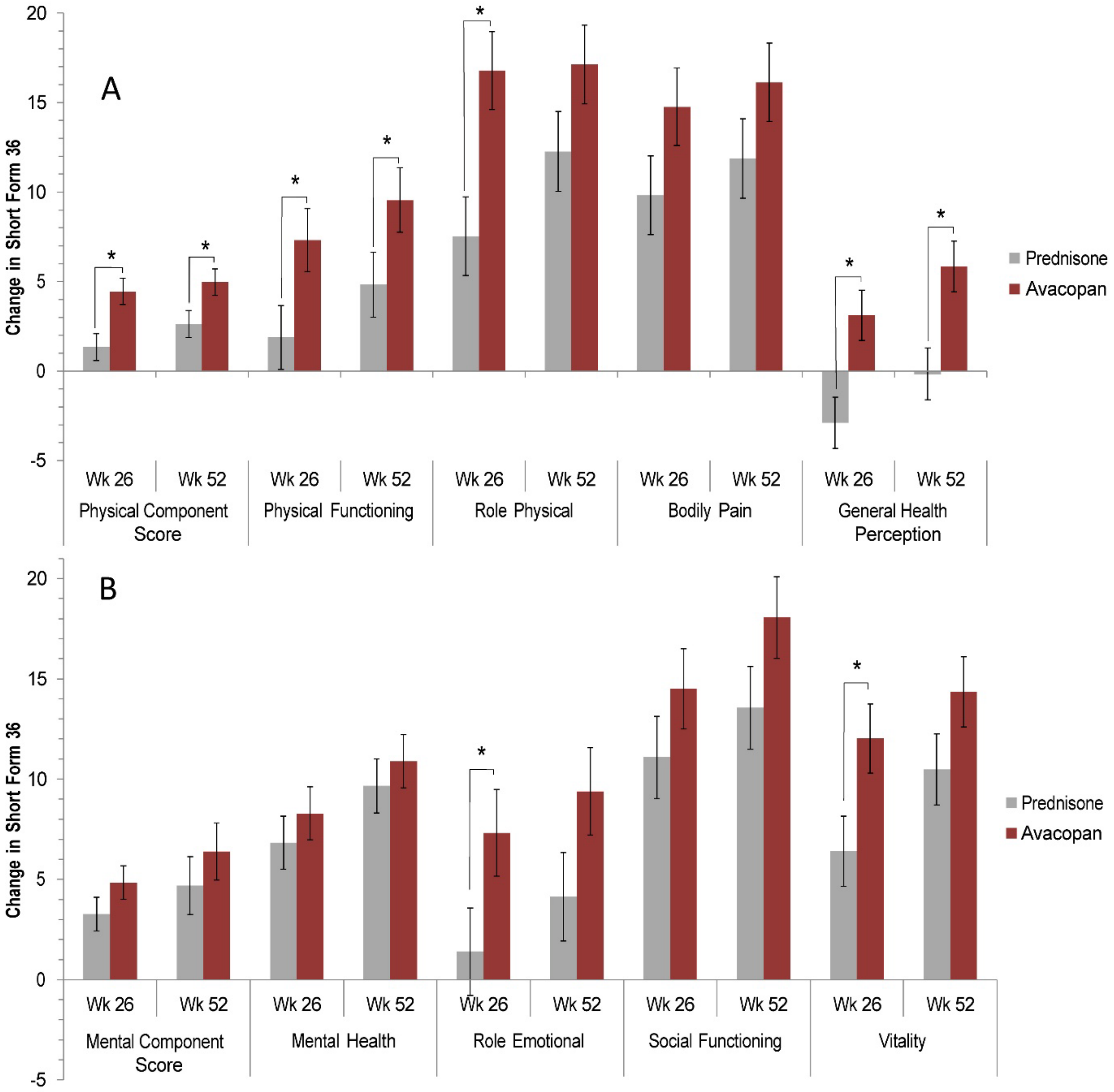

A. Change from baseline to week 26 and week 52 in health-related quality of life measurements based on the Medical Outcomes Study Short Form 36 version 2 Physical Component Score, individual physical component domains, and General Health Perception

B. Change from baseline to week 26 and week 52 in health-related quality of life measurements based on the Medical Outcomes Study Short Form 36 version 2 Mental Component Score, and individual mental component domains.

Data are presented as least squares means ± standard error of the means.

* Indicates where 95% confidence intervals of the differences between the avacopan and prednisone group do not include 0. The two treatment groups were compared by mixed effects model for repeated measures analysis with treatment group, study visit, treatment-by-visit interaction, and randomization stratum as factors and baseline as covariate.

**Figure S7. EQ-5D-5L**

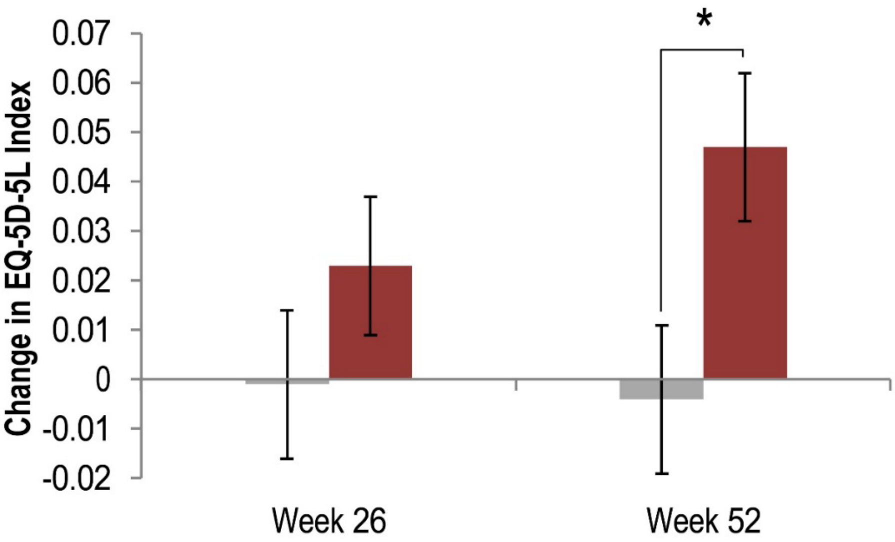

Change from baseline to week 26 and 52 in health-related quality of life measurement EQ-5D-5L index.

Data are presented as least squares means ± standard error of the means.

* Indicates where 95% confidence intervals of the differences between the avacopan and prednisone group do not include 0. The two treatment groups were compared by mixed effects model for repeated measures analysis with treatment group, study visit, treatment-by-visit interaction, and randomization stratum as factors and baseline as covariate.

#### 11. TABLES

**Table S1. Dose (mg/kg) for Oral or IV Cyclophosphamide Based on Age and Estimated Glomerular Filtration Rate**

| Age (years) | Oral Cyclophosphamide Dose (mg/kg/day) | | IV Cyclophosphamide Dose (mg/kg) | |
|---|---|---|---|---|
| | Estimated Glomerular Filtration Rate (mL/min/1.73 $m^2$) | | Estimated Glomerular Filtration Rate (mL/min/1.73 $m^2$) | |
| | >30 | ≤30 | >30 | ≤30 |
| <60 | 2 | 1.5 | 15 | 12.5 |
| 60-70 | 1.5 | 1.25 | 12.5 | 10 |
| >70 | 1.25 | 1 | 10 | 7.5 |

**Table S2. Pre-Specified Prednisone Dose Tapering Schedule for the Two Treatment Groups**

| Study Day | Avacopan Group | Prednisone Group | | | |
|---|---|---|---|---|---|
| | Daily Prednisone Dose* | | | | |
| | All† | Adults | | Adolescents | |
| | | ≥55 kg | <55 kg | >37 kg | ≤37 kg |
| Week 1 | 0 | 60 mg | 45 mg | 45 mg | 30 mg |
| Week 2 | 0 | 45 mg | 45 mg | 45 mg | 30 mg |
| Week 3 | 0 | 30 mg | 30 mg | 30 mg | 30 mg |
| Week 4 to 6 | 0 | 25 mg | 25 mg | 25 mg | 25 mg |
| Week 7 and 8 | 0 | 20 mg | 20 mg | 20 mg | 20 mg |
| Week 9 and 10 | 0 | 15 mg | 15 mg | 15 mg | 15 mg |
| Week 11 to 14 | 0 | 10 mg | 10 mg | 10 mg | 10 mg |
| Week 15 to 20 | 0 | 5 mg | 5 mg | 5 mg | 5 mg |
| ≥ Week 21 | 0 | 0 | 0 | 0 | 0 |

* Prednisone was supplied to study centers as 20 mg and 5 mg tablets, over-encapsulated with hard gelatin capsules to maintain the blind. Prednisone-matching placebo was given as gelatin capsules to the avacopan group. Dosing instructions for each period were provided to each study center.

† Any patients who were still taking 20 mg prednisone (non-study drug) on Day 1 were tapered to 0 mg by the end of week 4.

#### Table S3. Glucocorticoid Toxicity Index Version 2.0 Items with Scoring

| Feature/Body System | Item Weight |
| --- | --- |
| **Body Mass Index (BMI)** | |
| Decrease of ≥5 BMI units | –36 |
| Decrease of >2 but <5 BMI units | –21 |
| No significant change in BMI (±2 BMI units) | 0 |
| Increase of >2 to <5 BMI units | 21 |
| Increase of 5 or more BMI units | 36 |
| **Glucose tolerance** | |
| Improvement in HbA1c AND decrease in medication | –44 |
| Improvement in HbA1c OR decrease in medication | –32 |
| No significant change | 0 |
| Increase in HbA1c OR increase in medication | 32 |
| Increase in HbA1c AND increase in medication | 44 |
| **Blood pressure** | |
| Improvement in BP AND decrease in medication | –44 |
| Improvement in BP OR decrease in medication | –19 |
| No significant change in blood pressure | 0 |
| Increase in BP OR increase in medication | 19 |
| Increase in BP AND increase in medication | 44 |
| **Lipids** | |
| Decrease in LDL AND decrease in medication | –30 |
| Decrease in LDL OR decrease in medication | –10 |
| No significant change in lipids | 0 |
| Increase in LDL OR increase in medication | 10 |
| Increase in LDL AND increase in medication | 30 |
| **Steroid myopathy** | |
| Moderate weakness to none | –63 |
| Moderate to Mild weakness | –54 |
| Mild weakness to none | –9 |
| No significant change | 0 |
| None to mild weakness | 9 |
| Mild to moderate weakness | 54 |
| None to Moderate weakness | 63 |

| Feature/Body System | Item Weight |
|---|---|
| **Skin toxicity** | |
| Decrease in Skin Toxicity - Moderate to None | –26 |
| Decrease in Skin Toxicity - Moderate to Mild | –18 |
| Decrease in Skin Toxicity - Mild to None | –8 |
| No significant change | 0 |
| Increase in Skin Toxicity - None to Mild | 8 |
| Increase in Skin Toxicity - Mild to Moderate | 18 |
| Increase in Skin Toxicity - None to Moderate | 26 |
| **Neuropsychiatric (NP) toxicity** | |
| Decrease in NP Toxicity - Moderate to None | –74 |
| Decrease in NP Toxicity - Moderate to Mild | –63 |
| Decrease in NP Toxicity - Mild to None | –11 |
| No significant change | 0 |
| Increase in NP Toxicity - None to Mild | 11 |
| Increase in NP Toxicity – Mild to Moderate | 63 |
| Increase in NP Toxicity - None to Moderate | 74 |
| **Infection** | |
| No significant infection | 0 |
| Oral/vaginal candidiasis or uncomplicated zoster | 19 |
| Grade 3, 4 or 5 infection | 93 |

BMI = body mass index; BP = blood pressure; HbA1c = hemoglobin A1c; LDL = low density lipoprotein; NP = neuropsychiatric

**Table S4. Remission at Week 26 and Sustained Remission at Week 52 for the Per-Protocol Population – 2026 Readjudication***

| | Prednisone (N=161) | Avacopan (N=162) | P-value for Difference Between Groups[†] |
|---|---|---|---|
| Remission[‡] at week 26—no. (%) | 110 (68.3) | 112 (69.1) | <0.0001 (non-inferiority)<br>0.35 (superiority) |
| Estimate of common difference in percentages | -- | 1.9 | |
| Two-sided 95% confidence interval for common difference | -- | –7.8, 11.7 | |
| Sustained remission[§] at week 52—no. (%) | 86 (53.4) | 101 (62.3) | <0.0001 (non-inferiority)<br>0.0311 (superiority) |
| Estimate of common difference in percentages | -- | 9.7 | |
| Two-sided 95% confidence interval for common difference | -- | –0.5, 19.8 | |

* Patients who had major protocol deviations regarding inclusion criteria or who had missing week 26 data were excluded from the Per-Protocol population. Additionally, those who were not compliant with taking avacopan/placebo (based on returned capsule counts), and those who used non-protocol specified medications were imputed as non-remitters at week 26 or 52, as applicable.
Subjects who received low dose oral glucocorticoids (≤10 mg/day) for treatment of adrenal insufficiency are classified as responders for purposes of assessment of the primary endpoint if all other criteria for response are met.
† One-sided P-values
‡ Remission was defined as having a BVAS of 0 at week 26 and not having received any glucocorticoids for vasculitis within the 4 weeks prior to the week 26 visit.
§ Sustained remission at week 52 is based on assessment by the blinded Adjudication Committee and was defined as remission at week 26 (BVAS of 0 and not taking glucocorticoids for treatment of vasculitis within 4 weeks prior to week 26) and remission at week 52 (BVAS of 0 and not taking glucocorticoids for treatment of vasculitis within 4 weeks prior to Week 52) and without relapse between week 26 and 52.

**Table S5. Remission at Week 26 for Each Subgroup- 2019 Readjudication ***

| | Prednisone (N=164) | Avacopan (N=166) |
|---|---|---|
| All Patients* | 115 / 164 (70.1%) | 120 / 166 (72.3%) |
| Disease Status | | |
| Newly diagnosed patients | 76 / 114 (66.7%) | 76 / 115 (66.1%) |
| Relapsing disease | 39 / 50 (78.0%) | 44 / 51 (86.3%) |
| ANCA Type | | |
| Anti-proteinase 3 positive | 50 / 70 (71.4%) | 51 / 72 (70.8%) |
| Anti-myeloperoxidase positive | 65 / 94 (69.1%) | 69 / 94 (73.4%) |
| Background Treatment | | |
| Cyclophosphamide | 34 / 57 (59.6%) | 37 / 59 (62.7%) |
| Rituximab | 81 / 107 (75.7%) | 83 / 107 (77.6%) |
| Type of ANCA-Associated Vasculitis | | |
| Granulomatosis with polyangiitis | 65 / 90 (72.2%) | 65 / 91 (71.4%) |
| Microscopic polyangiitis | 50 / 74 (67.6%) | 55 / 75 (73.3%) |

* Results are shown for n / N (%), where n = the number of remitters and N = the number of patients in each stratum. Remission at week 26 was defined as achieving a BVAS of zero and not having received any glucocorticoid treatment for vasculitis within 4 weeks prior to the week 26 visit.

#### Table S6. Sustained Remission at Week 52 for Each Subgroup- 2019 Adjudication*

| | Prednisone (N=164) | Avacopan (N=166) |
|---|---|---|
| All Patients* | 90 / 164 (54.9%) | 109 / 166 (65.7%) |
| Disease Status | | |
| Newly diagnosed patients | 66 / 114 (57.9%) | 70 / 115 (60.9%) |
| Relapsing disease | 24 / 50 (48.0%) | 39 / 51 (76.5%) |
| ANCA Type | | |
| Anti-proteinase 3 positive | 40 / 70 (57.1%) | 43 / 72 (59.7%) |
| Anti-myeloperoxidase positive | 50 / 94 (53.2%) | 66 / 94 (70.2%) |
| Background Treatment | | |
| Cyclophosphamide | 30 / 57 (52.6%) | 33 / 59 (55.9%) |
| Rituximab | 60 / 107 (56.1%) | 76 / 107 (71.0%) |
| Type of ANCA-Associated Vasculitis | | |
| Granulomatosis with polyangiitis | 52 / 90 (57.8%) | 56 / 91 (61.5%) |
| Microscopic polyangiitis | 38 / 74 (51.4%) | 53 / 75 (70.7%) |

*Results are shown for n / N (%), where n = the number of remitters and N = the number of patients in each stratum. Sustained remission was defined as remission at week 26 and remission at week 52 (BVAS of 0 and not taking glucocorticoids for treatment of vasculitis within 4 weeks prior to Week 52) and without relapse between week 26 and 52.

**Table S7: Week 26 and Week 52 Adjudication Outcomes for the 9 Participants for Whom Procedures Used to Assess the Primary Outcome in 2019 Raised Concern.**

| | Treatment Arm | Week 26 | | Week 52 | |
|---|---|---|---|---|---|
| | | 2019 adjudication | 2026 adjudication | 2019 adjudication | 2026 adjudication |
| 1 | Prednisone | In remission | In remission | Not in remission | Not in remission |
| 2 | Avacopan | In remission | Not in remission | In remission | Not in remission |
| 3 | Avacopan | In remission | In remission | In remission | In remission |
| 4 | Avacopan | In remission | In remission | In remission | In remission |
| 5 | Avacopan | In remission | In remission | In remission | In remission |
| 6 | Avacopan | In remission | In remission | In remission | In remission |
| 7 | Prednisone | In remission | In remission | Not in remission | Not in remission |
| 8 | Avacopan | In remission | Not in remission | Not in remission | Not in remission |
| 9 | Prednisone | In remission | In remission | Not in remission | Not in remission |

#### Table S8. All Secondary End Point Results

| | Prednisone (N=164) | Avacopan (N=166) | Difference Avacopan minus Prednisone (95% Confidence Interval) |
|---|---|---|---|
| Glucocorticoid Toxicity Index Cumulative Worsening Score | | | |
| Week 13 (LSM ± SEM) | 36.6 ± 3.41 (n=161) | 25.7 ± 3.40 (n=160) | –11.0 (–19.7 to –2.2) |
| Week 26 (LSM ± SEM) | 56.6 ± 3.45 (n=153) | 39.7 ± 3.43 (n=154) | –16.8 (–25.6 to –8.0) |
| Glucocorticoid Toxicity Index Aggregate Improvement Score | | | |
| Week 13 (LSM ± SEM) | 23.2 ± 3.46 (n=161) | 9.9 ± 3.45 (n=160) | –13.3 (–22.2 to –4.4) |
| Week 26 (LSM ± SEM) | 23.4 ± 3.50 (n=153) | 11.2 ± 3.48 (n=154) | –12.1 (–21.1 to –3.2) |
| Estimated Glomerular Filtration Rate (mL/min/1.73 $m^2$) in patients with renal disease at baseline based on Birmingham Vasculitis Activity Score | | | |
| Baseline (mean ± SEM) | 45.6 ± 2.36 (n=134) | 44.6 ± 2.42 (n=131) | |
| Change from baseline to week 26 (LSM ± SEM) | 2.9 ± 1.03 (n=127) | 5.8 ± 1.04 (n=121) | 2.9 (0.1 to 5.8) |
| Change from baseline to week 52 (LSM ± SEM) | 4.1 ± 1.03 (n=125) | 7.3 ± 1.05 (n=119) | 3.2 (0.3 to 6.1) |
| Health-Related Quality of Life, Short Form 36 version 2.0, Physical Component Score | | | |
| Baseline (mean ± SEM) | 40.1 ± 0.83 (n=160) | 39.2 ± 0.80 (n=165) | |
| Change from baseline to week 26 (LSM ± SEM) | 1.34 ± 0.74 (n=147) | 4.45 ± 0.73 (n=153) | 3.10 (1.17 to 5.03) |
| Change from baseline to week 52 (LSM ± SEM) | 2.63 ± 0.75 (n=144) | 4.98 ± 0.74 (n=147) | 2.35 (0.40 to 4.31) |
| Health-Related Quality of Life, Short Form 36 version 2.0, Mental Component Score | | | |
| Baseline (mean ± SEM) | 42.1 ± 1.05 (n=160) | 44.2 ± 0.98 (n=166) | |
| Change from baseline to week 26 (LSM ± SEM) | 3.27 ± 0.84 (n=147) | 4.85 ± 0.83 (n=154) | 1.58 (–0.61 to 3.77) |
| Change from baseline to week 52 (LSM ± SEM) | 4.69 ± 0.85 (n=144) | 6.39 ± 0.84 (n=148) | 1.70 (–0.52 to 3.92) |

| | Prednisone (N=164) | Avacopan (N=166) | Difference Avacopan minus Prednisone (95% Confidence Interval) |
|---|---|---|---|
| Health-Related Quality of Life, EQ-5D-5L, Visual Analogue Scale | | | |
| Baseline (mean ± SEM) | 63.4 ± 1.78 (n=162) | 65.8 ± 1.51 (n=166) | |
| Change from baseline to week 26 (LSM ± SEM) | 5.5 ± 1.39 (n=150) | 9.1 ± 1.38 (n=153) | 3.6 (–0.1 to 7.2) |
| Change from baseline to week 52 (LSM ± SEM) | 7.1 ± 1.41 (n=146) | 13.0 ± 1.39 (n=149) | 5.9 (2.3 to 9.6) |
| Health-Related Quality of Life, EQ-5D-5L, Index | | | |
| Baseline (mean ± SEM) | 0.77 ± 0.018 (n=160) | 0.75 ± 0.018 (n=166) | |
| Change from baseline to week 26 (LSM ± SEM) | 0.00 ± 0.015 (n=146) | 0.02 ± 0.014 (n=152) | 0.02 (–0.01 to 0.06) |
| Change from baseline to week 52 (LSM ± SEM) | 0.00 ± 0.015 (n=145) | 0.05 ± 0.015 (n=149) | 0.05 (0.01 to 0.09) |
| Birmingham Vasculitis Activity Score = 0 at week 4 | 113 (68.9) | 104 (62.7) | –5.6 (–15.4 to 4.2) |
| Relapses* during the treatment period after remission has been achieved at week 26 (2019 Adjudication) | 14 of 115 (12.2) | 9 of 120 (7.5) | –6.0 (–14.4 to 2.4) |
| Relapses* during the treatment period after remission has been achieved at week 26 (2026 Re-adjudication) | 12 of 110 (10.9) | 7 of 113 (6.2) | –6.0 (–14.6 to 2.6) |
| Urinary Albumin:Creatinine Ratio in Patients with Renal Disease (based on BVAS) and UACR of at least 10 mg/g creatinine at baseline† | | | |
| Baseline (geometric mean, range), mg/g | 312.2 (11-5367) (n=128) | 432.9 (20-6461) (n=125) | |
| Percent change from baseline to week 4 (LSM ± SEM) | 0 ± 9.3) (n=124) | –40 ± 9.5 (n=121) | –40 (–53 to –22) |
| Percent change from baseline to week 13 (LSM ± SEM) | –49 ± 9.4 (n=121) | –55 ± 9.6 (n=116) | –12 (–32 to 13) |
| Percent change from baseline to week 26 (LSM ± SEM) | –70 ± 9.5 (n=118) | –63 ± 9.7 (n=113) | 25 (–3 to 61) |
| Percent change from baseline to week 52 (LSM ± SEM) | –77 ± 9.6 (n=114) | –74 ± 9.8 (n=109) | 12 (–14 to 45) |

| | Prednisone (N=164) | Avacopan (N=166) | Difference Avacopan minus Prednisone (95% Confidence Interval) |
|---|---|---|---|
| Urinary Monocyte Chemoattractant Protein-1:Creatinine Ratio in Patients with Renal Disease (based on BVAS) at baseline† | | | |
| Baseline (geometric mean, range), pg/mg | 947.8 (160-6525) (n=130) | 983.8 (138-6145) (n=127) | |
| Percent change from baseline to week 4 (LSM ± SEM) | –22 ± 5.6 (n=124) | –25 ± 5.7 (n=120) | –4 (–17 to 12) |
| Percent change from baseline to week 13 (LSM ± SEM) | –52 ± 5.7 (n=120) | –59 ± 5.8 (n=113) | –15 (–28 to –1) |
| Percent change from baseline to week 26 (LSM ± SEM) | –64 ± 5.7 (n=117) | –67 ± 5.9 (n=106) | –9 (–22 to 7) |
| Percent change from baseline to week 52 (LSM ± SEM) | –71 ± 5.9 (n=108) | –73 ± 6.0 (n=106) | –10 (–23 to 6) |
| Vasculitis Damage Index | | | |
| Baseline (mean ± SEM) | 0.72 ± 0.109 (n=163) | 0.66 ± 0.120 (n=165) | |
| Change from baseline to week 26 (LSM ± SEM) | 0.97 ± 0.092 (n=155) | 1.06 ± 0.090 (n=161) | 0.09 (–0.14 to 0.32) |
| Change from baseline to week 52 (LSM ± SEM) | 1.15 ± 0.093 (n=151) | 1.17 ± 0.091 (n=150) | 0.02 (–0.21 to 0.25) |

LSM = least squares mean; SEM = standard error of the mean

* The assessment of remission at week 26 and the relapse after week 26 are both based on the assessment by the Adjudication Committee.

A relapse was defined as worsening of disease, after having previously achieved remission (BVAS = 0), that involves:

- one or more major item in the BVAS, or
- three or more minor items in the BVAS, or
- one or two minor items in the BVAS recorded at two consecutive study visits.

† Urinary albumin:creatinine ratio and urinary monocyte chemoattractant protein-1:creatinine ratio percentage change from baseline are based on ratios of geometric means of visit over baseline.

**Table S9. Glucocorticoid Use (from All Sources) During the 52-Week Treatment Period of the Study**

| | **Prednisone (N=164)** | **Avacopan (N=166)** |
|---|---|---|
| Any Oral or IV Use (mg)* | | |
| n (%) | 164 (100.0%) | 145 (87.3%) |
| total dose, mean ± SD | 3846.9 ± 1912.90 | 1675.5 ± 2812.55 |
| daily dose, mean ± SD | 12.5 ± 9.95 | 5.4 ± 9.08 |
| total dose, median (Min, Max) | 3097.5 (760, 13383) | 600.0 (0, 21680) |
| Oral Prednisone Study Medication (mg) | | |
| n (%) | 164 (100.0%) | Not applicable |
| total dose, mean ± SD | 2389.2 ± 624.31 | Not applicable |
| daily dose, mean ± SD | 7.8 ± 6.11 | Not applicable |
| Oral, Other Than Prednisone Study Medication (mg)† | | |
| n (%) | 115 (70.1%) | 112 (67.5%) |
| total dose, mean ± SD | 903.9 ± 1327.85 | 964.9 ± 1979.27 |
| daily dose, mean ± SD | 2.7 ± 3.96 | 3.1 ± 5.83 |
| Any IV Use (mg) | | |
| n (%) | 124 (75.6%) | 120 (72.3%) |
| total dose, mean ± SD | 553.8 ± 844.48 | 710.5 ± 1241.51 |
| daily dose, mean ± SD | 2.0 ± 4.10 | 2.4 ± 4.98 |

n = number of patients in the specific category; N = number of patients in the treatment group; % = n/N x 100
*Prednisone-equivalent dose calculated across all patients in the treatment group.
†Sources of oral glucocorticoids include the initial 4-week taper, glucocorticoid use for non-vasculitis reasons such as adrenal insufficiency, and limited use for minor worsening or persistence of vasculitis.

**Table S10. Glucocorticoid Use and Other Immunosuppressant Use (Other Than Protocol-Specified) by Study Period**

| | Prednisone (N=164) | Avacopan (N=166) |
|---|---|---|
| Glucocorticoid use | | |
| Screening Period (Week –2 to Week –1) | | |
| Any Oral or IV (mg)* | | |
| n (%) | 135 (82.3%) | 125 (75.3%) |
| total dose, mean ± SD | 978.0 ± 1157.49 | 907.3 ± 1145.91 |
| daily dose, mean ± SD | 69.9 ± 82.68 | 64.8 ± 81.85 |
| Oral (mg) | | |
| n (%) | 113 (68.9%) | 99 (59.6%) |
| total dose, mean ± SD | 219.3 ± 256.26 | 198.3 ± 277.58 |
| daily dose, mean ± SD | 15.7 ± 18.30 | 14.2 ± 19.83 |
| IV (mg) | | |
| n (%) | 73 (44.5%) | 63 (38.0%) |
| total dose, mean ± SD | 758.8 ± 1110.97 | 709.1 ± 1131.44 |
| daily dose, mean ± SD | 54.2 ± 79.36 | 50.6 ± 80.82 |
| Week 1 to Week 4 | | |
| Any Oral or IV (mg)* | | |
| n (%) | 164 (100.0%) | 138 (83.1%) |
| total dose, mean ± SD | 1649.7 ± 541.49 | 625.4 ± 1164.09 |
| daily dose, mean ± SD | 57.2 ± 18.57 | 21.6 ± 40.13 |
| Prednisone Study Medication (mg) | | |
| n (%) | 164 (100.0%) | Not applicable |
| total dose, mean ± SD | 1148.3 ± 168.22 | Not applicable |
| daily dose, mean ± SD | 39.8 ± 5.81 | Not applicable |
| Oral, Other Than Prednisone Study Medication (mg) | | |
| n (%) | 89 (54.3%) | 93 (56.0%) |
| total dose, mean ± SD | 155.5 ± 251.53 | 214.8 ± 689.55 |
| daily dose, mean ± SD | 5.4 ± 8.67 | 7.4 ± 23.78 |
| IV (mg) | | |
| n (%)† | 112 (68.3%) | 111 (66.9%) |
| total dose, mean ± SD | 345.9 ± 454.81 | 410.6 ± 628.93 |
| daily dose, mean ± SD | 12.0 ± 15.70 | 14.2 ± 21.68 |

| | **Prednisone (N=164)** | **Avacopan (N=166)** |
|---|---|---|
| Week 5 to Week 26 | | |
| Any Oral or IV (mg)* | | |
| n (%) | 161 (98.2%) | 52 (31.3%) |
| total dose, mean ± SD | 1746.4 ± 1172.13 | 771.6 ± 1808.54 |
| daily dose, mean ± SD | 11.5 ± 7.75 | 5.4 ± 12.75 |
| Prednisone Study Medication (mg) | | |
| n (%) | 157 (95.7%) | Not applicable |
| total dose, mean ± SD | 1264.1 ± 495.98 | Not applicable |
| daily dose, mean ± SD | 8.4 ± 3.32 | Not applicable |
| Oral, Other Than Prednisone Study Medication (mg) | | |
| n (%) | 53 (32.3%) | 48 (28.9%) |
| total dose, mean ± SD | 336.3 ± 708.90 | 500.8 ± 1146.39 |
| daily dose, mean ± SD | 2.2 ± 4.68 | 3.4 ± 7.54 |
| IV (mg) | | |
| n (%) | 21 (12.8%) | 21 (12.7%) |
| total dose, mean ± SD | 146.0 ± 597.87 | 270.8 ± 1036.98 |
| daily dose, mean ± SD | 1.0 ± 3.89 | 2.0 ± 8.04 |
| Week 27 to End of Treatment | | |
| Any Oral or IV (mg)* | | |
| n (%) | 64 (39.0%) | 45 (27.1%) |
| total dose, mean ± SD | 511.0 ± 962.32 | 323.1 ± 743.63 |
| daily dose, mean ± SD | 3.0 ± 6.15 | 1.8 ± 4.24 |
| Oral (mg) | | |
| n (%) | 58 (35.4%) | 40 (24.1%) |
| total dose, mean ± SD | 442.7 ± 775.33 | 283.2 ± 642.48 |
| daily dose, mean ± SD | 2.6 ± 5.09 | 1.6 ± 3.72 |
| IV (mg) | | |
| n (%) | 19 (11.6%) | 14 (8.4%) |
| total dose, mean ± SD | 68.4 ± 340.09 | 40.2 ± 210.35 |
| daily dose, mean ± SD | 0.4 ± 1.91 | 0.2 ± 1.15 |

| | Prednisone (N=164) | Avacopan (N=166) |
|---|---|---|
| Non-protocol specified immunosuppressant use‡ | | |
| Week 1 to Week 26, n (%) | | |
| Any Immunosuppressant | 16 (9.8%) | 14 (8.4%) |
| Rituximab | 11 (6.7%) | 6 (3.6%) |
| Cyclophosphamide | 3 (1.8%) | 3 (1.8%) |
| Azathioprine | 1 (0.6%) | 5 (3.0%) |
| Methotrexate/Methotrexate Sodium | 0 (0.0%) | 0 (0.0%) |
| Mycophenolate Mofetil/ Mycophenolic Acid | 1 (0.6%) | 0 (0.0%) |
| Week 27 to End of Treatment, n (%) | | |
| Any Immunosuppressant | 25 (15.2%) | 19 (11.4%) |
| Rituximab | 22 (13.4%) | 15 (9.0%) |
| Cyclophosphamide | 1 (0.6%) | 3 (1.8%) |
| Azathioprine | 0 (0.0%) | 0 (0.0%) |
| Methotrexate/Methotrexate Sodium | 2 (1.2%) | 1 (0.6%) |
| Mycophenolate Mofetil/ Mycophenolic Acid | 0 (0.0%) | 1 (0.6%) |
| Week 1 to End of Treatment, n (%) | | |
| Any Immunosuppressant | 36 (22.0%) | 29 (17.5%) |
| Rituximab | 30 (18.3%) | 19 (11.4%) |
| Cyclophosphamide | 3 (1.8%) | 6 (3.6%) |
| Azathioprine | 1 (0.6%) | 5 (3.0%) |
| Methotrexate/Methotrexate Sodium | 2 (1.2%) | 1 (0.6%) |
| Mycophenolate Mofetil/ Mycophenolic Acid | 1 (0.6%) | 1 (0.6%) |

n = number of patients in the specific category; N = number of patients in the treatment group; % = n/N x 100
* Prednisone-equivalent dose calculated across all patients in the treatment group.
† Includes IV glucocorticoids as pre-medication for rituximab or cyclophosphamide. A total of 19 patients in the prednisone group and 21 in the avacopan group received IV glucocorticoids that were not pre-medication.
‡ Immunosuppressant use for worsening of vasculitis or relapses

**Table S11. All Serious Infections by Treatment Group**

| | Prednisone (N=164) | Avacopan (N=166) |
|---|---|---|
| Any serious infection* | 25 (15.2%) | 22 (13.3%) |
| Pneumonia† | 9 (5.5%) | 9 (5.4%) |
| Urinary tract infection | 2 (1.2%) | 3 (1.8%) |
| Device related infection | 0 (0.0%) | 2 (1.2%) |
| Influenza | 1 (0.6%) | 2 (1.2%) |
| Bronchitis | 1 (0.6%) | 1 (0.6%) |
| Campylobacter gastroenteritis | 0 (0.0%) | 1 (0.6%) |
| Hepatitis B reactivation‡ | 0 (0.0%) | 1 (0.6%) |
| Infective exacerbation of chronic obstructive pulmonary disease | 0 (0.0%) | 1 (0.6%) |
| Neutropenic sepsis | 0 (0.0%) | 1 (0.6%) |
| Post procedural sepsis | 0 (0.0%) | 1 (0.6%) |
| Sepsis | 1 (0.6%) | 1 (0.6%) |
| Urosepsis | 0 (0.0%) | 1 (0.6%) |
| Aspergillus infection | 1 (0.6%) | 0 (0.0%) |
| Bacteremia | 1 (0.6%) | 0 (0.0%) |
| Cryptococcosis | 1 (0.6%) | 0 (0.0%) |
| Fungal infection | 1 (0.6%) | 0 (0.0%) |
| Herpes zoster | 2 (1.2%) | 0 (0.0%) |
| Infectious pleural infusion | 2 (1.2%) | 0 (0.0%) |
| Meningitis | 1 (0.6%) | 0 (0.0%) |
| Ophthalmic herpes simplex | 1 (0.6%) | 0 (0.0%) |
| Parainfluenza virus infection | 1 (0.6%) | 0 (0.0%) |
| Respiratory syncytial virus infection | 2 (1.2%) | 0 (0.0%) |
| Respiratory tract infection viral | 1 (0.6%) | 0 (0.0%) |
| Staphylococcal infection | 1 (0.6%) | 0 (0.0%) |
| Subcutaneous abscess | 1 (0.6%) | 0 (0.0%) |

*A total of 15 patients (9.1%) in the prednisone group and 13 patients (7.8%) in the avacopan group had serious infections during the first 20 weeks of the study. A total of 10 patients (6.1%) in the prednisone group and 9 patients (5.4%) in the avacopan group had serious infections from week 21 through the end of the study.

† Includes adverse event terms of pneumonia, pneumonia bacterial, atypical pneumonia, pneumonia cytomegaloviral, pneumonia haemophilus, and lower respiratory tract infection.

‡ One patient had hepatitis B reactivation during the 8-week study drug free follow-up period. The patient had two rituximab infusions prior to the event.

**Table S12. Prospectively Identified System Organ Clusters and MedDRA Preferred Terms[15] for Adverse Events Potentially Associated with Glucocorticoid Toxicity**

| System Organ Cluster | Included MedDRA Preferred Terms |
|---|---|
| Cardiovascular | Oedema peripheral, peripheral swelling, oedema, hypertension, arteriosclerosis, hypertensive emergency, hypercholesterolaemia, dyslipidaemia, hypertriglyceridaemia, hypervolaemia, fluid retention, hyperlipidaemia, angina pectoris, cardiac failure, acute myocardial infarction, cardiovascular insufficiency, congestive cardiomyopathy, myocardial ischemia, myocardial infarction, blood pressure increased, blood cholesterol increased, low density lipoprotein increased |
| Infections | All serious infections |
| Gastrointestinal | Pancreatitis acute, duodenal ulcer, gastritis erosive, gastritis, gastrointestinal disorder, pancreatitis |
| Psychological | Insomnia, depression, anxiety, depressed mood, mania, affective disorder, agitation, libido decreased, nervousness, confusional state, irritability, major depression, mental status changes, mood altered, poor quality sleep |
| Endocrine/metabolic | Hyperglycaemia, hypokalaemia, diabetes mellitus, type 2 diabetes mellitus, central obesity, diabetes mellitus inadequate control, glucose tolerance impaired, blood potassium decreased, waste circumference increased, weight increased, blood glucose increased, Cushingoid, adrenal insufficiency, Cushing's syndrome, menorrhagia, metrorrhagia, gynaecomastia, influenza like illness, systemic inflammatory response syndrome |
| Dermatological | Dermatitis acneiform, acne, ecchymosis, hirsutism, skin atrophy, skin striae, increased tendency to bruise |
| Musculoskeletal | Myopathy, osteopenia, osteoporosis, osteonecrosis, muscle atrophy, muscular weakness, hip fracture, humerus fracture, lower limb fracture, tendon rupture, wrist fracture, lumbar vertebral fracture, spinal compression fracture |
| Ophthalmological | Cataract, retinopathy hypertensive, cataract nuclear, glaucoma, open angle glaucoma, intraocular pressure increased |

MedDRA = Medical Dictionary for Regulatory Activities
